## Supplementary material for "Monitoring and responding to emerging infectious diseases in a university setting: A case study using COVID-19": S2 Supporting Information: betaAndRateOfImportationCalculations.R

```
#This script calculates the base probabilities of transmission and the rates of imported infections that 
#   were used in Soda, Chen, Feinn, and Hill. See S1 Appendix for an explanation of these calculations.
#Author: K. James Soda
#Date  : Jan. 10, 2023

#This code estimates the probability that a symptomatic individual leads to a reported case on any
#   specific day and stores this value as result.
geometricRoot <- function(p,duration,totalProb){
	return(pgeom(duration,p) - totalProb)
}

result = uniroot(geometricRoot,c(10**-5,1),duration = 17,totalProb = 1/3,
	tol = .Machine$double.eps^0.5)


##This function will estimate the rate of imported infections for a given student population size based 
##  on a four-day time series of case incidences in the surrounding community.
##INPUTS:
#incidence  = vector (numeric); reported cases (out of 100000) over a four-day period. Each element
#		  is the number of cases in one day.
#p_detect   = numeric; probability that a symptomatic infection will lead to a reported case on any given 
#             day
#p_symp     = numeric; probability that an infection will become symptomatic
#population = int; population size for the surrounding community
#nStudent   = int; size of the student population
#nContacts  = numeric; average number of contacts a student has per day with individuals in the 
#             surrounding community.
##OUTPUT
#data frame; the data frame has three columns:
#    daysToSymp = integer; the assumed number of days between when an exposed individual becomes
#                 infectious and when they potentially develop symptoms. There are two entries,
#                 one day and three days. Only the three day results were used in the study.
#    prevalence = numeric; estimated prevalence in the community as the proportion of individuals in the
#                 surrounding community who are infectious. 
#    importRate = numeric; the estimated rate of imported infections in infections/week.

estimateImports <- function(incidence,
				    p_detect,
				    p_symp = 0.6,
			          population = 3565287,
				    nStudents = 3636,
				    nContacts = 1,
				    probTransmission = 0.016){
	#Convert the incidence to raw number of cases
	incidence = incidence * population/100000

	#Estimate the number of symptomatic infections
	n_symp = incidence[1] / p_detect - 1

	#Add the asymptomatic cases
	n_post = n_symp * (1 + ((1 - p_symp)/p_symp))

	#Add the pre-symptomatic but still infectious
	#Assuming 1 day
	n_pre1 = incidence[2] * (1 + (1 - p_symp)/p_symp)

	#Assuming 3 days
	n_pre3 = sum(incidence[2:4]) * (1 + (1 - p_symp)/p_symp)

	#Calculate the prevalences
	prev1 = round(n_post + n_pre1)/population
	prev3 = round(n_post + n_pre3)/population

	#Calculate the importation rates (in imported cases/week)
	import1 = log(nContacts) + log(nStudents) + log(prev1) + log(probTransmission)
	import1 = 7 * exp(import1)

	import3 = log(nContacts) + log(nStudents) + log(prev3) + log(probTransmission)
	import3 = 7 * exp(import3)


	#Provide the outcome
	return(data.frame(daysToSymp = c(1,3),prevalence = c(prev1,prev3),importRate = c(import1,import3)))
}


#This function calculates the base probability of transmission for a desired R_0
##INPUTS
#r0         = numeric; the desired R_0
#n_contacts = integer;  expected number of contacts an individual in the population is assumed to
#             have.
##OUTPUT
#numeric; the probability of  transmission that corresponds to the desired R_0

findBeta <- function(r0,n_contacts){
	#This age distribution for US citizens is based on data for 2018 on data.census.gov
      #NOTE: There is a slight discrepancy with the dataset above. See S1 Appendix.
	ageDistr = c(0.121,0.131,0.525,0.1095,0.1135)

	#These are the default age-specific susceptibilities for covasim
	#Note that age bins have been fused when they have the same susceptibilities
	sus = c(0.34,0.67,1,1.24,1.47)

	return(r0/(n_contacts * sum(ageDistr*sus) * 11/1.3))
}

##Rate of Imported Infections Based on Aug. 30 Case Incidence Data (all CT)
##NOTE: The probability of transmission is rounded to the number of decimal places used in the simulations.
#Average Contacts: 8, R_0: 1.5
#Residential Students (0.5 Contact)
estimateImports(c(3.5,3.9,3.9,3.6),result$root,nContacts = 0.5,nStudents = 3636,probTransmission = round(findBeta(1.5,8),6))
#Non-Residential Students (5.5 Contact)
estimateImports(c(3.5,3.9,3.9,3.6),result$root,nContacts = 5.5,nStudents = 3789,probTransmission = round(findBeta(1.5,8),6))


#Average Contacts: 8, R_0: 2.5
#Residential Students (0.5 Contact)
estimateImports(c(3.5,3.9,3.9,3.6),result$root,nContacts = 0.5,nStudents = 3636,probTransmission = round(findBeta(2.5,8),6))
#Non-Residential Students (5.5 Contact)
estimateImports(c(3.5,3.9,3.9,3.6),result$root,nContacts = 5.5,nStudents = 3789,probTransmission = round(findBeta(2.5,8),6))


#Average Contacts: 10, R_0: 1.5
#Residential Students (1 Contact)
estimateImports(c(3.5,3.9,3.9,3.6),result$root,nContacts = 1,nStudents = 3636,probTransmission = round(findBeta(1.5,10),7))
#Non-Residential Students (6 Contact)
estimateImports(c(3.5,3.9,3.9,3.6),result$root,nContacts = 6,nStudents = 3789,probTransmission = round(findBeta(1.5,10),7))


#Average Contacts: 10, R_0: 2.5
#Residential Students (1 Contact)
estimateImports(c(3.5,3.9,3.9,3.6),result$root,nContacts = 1,nStudents = 3636,probTransmission = round(findBeta(2.5,10),6))
#Non-Residential Students (6 Contact)
estimateImports(c(3.5,3.9,3.9,3.6),result$root,nContacts = 6,nStudents = 3789,probTransmission = round(findBeta(2.5,10),6))
```
