## Supplementary material for "Monitoring and responding to emerging infectious diseases in a university setting: A case study using COVID-19": S2 Supporting Information: estimateScalingRelationship.py

```
'''
This script estimates the proportional scaling relationship between the cumulative infections and the rate of imported infections under the four main
scenarios in Soda, Chen, Feinn, and Hill.
Author: K. James Soda
Date  : Jan. 9, 2023
'''

from datetime import datetime
import pandas as pd
import numpy as np
import scipy.optimize as opt


def slopeError(x,predictor,output):
    return sum((output - x * predictor)**2)

if __name__ == "__main__":
    #Import the data for R0 = 1.5, Contacts = 8
    infectTS_r15c8_half   = pd.read_csv(".\\sensitivityFiles\\cumInfectSeries_sc12_halfImport_r01.5_contact8c.csv",index_col = 0)
    infectTS_r15c8_full   = pd.read_csv(".\\sensitivityFiles\\cumInfectSeries_sc12_r01.5_contact8.csv",index_col = 0)
    infectTS_r15c8_double = pd.read_csv(".\\sensitivityFiles\\cumInfectSeries_sc12_doubleImport_r01.5_contact8c.csv",index_col = 0)

    #Import the data for R0 = 1.5, Contacts = 10
    infectTS_r15c10_half   = pd.read_csv(".\\sensitivityFiles\\cumInfectSeries_sc12_halfImport_r01.5_contact10c.csv",index_col = 0)
    infectTS_r15c10_full   = pd.read_csv(".\\sensitivityFiles\\cumInfectSeries_sc12_r01.5_contact10.csv",index_col = 0)
    infectTS_r15c10_double = pd.read_csv(".\\sensitivityFiles\\cumInfectSeries_sc12_doubleImport_r01.5_contact10c.csv",index_col = 0)

    #Import the data for R0 = 2.5, Contacts = 8
    infectTS_r25c8_half   = pd.read_csv(".\\sensitivityFiles\\cumInfectSeries_sc12_halfImport_r02.5_contact8c.csv",index_col = 0)
    infectTS_r25c8_full   = pd.read_csv(".\\sensitivityFiles\\cumInfectSeries_sc12_r02.5_contact8.csv",index_col = 0)
    infectTS_r25c8_double = pd.read_csv(".\\sensitivityFiles\\cumInfectSeries_sc12_doubleImport_r02.5_contact8c.csv",index_col = 0)

    #Import the data for R0 = 2.5, Contacts = 10
    infectTS_r25c10_half   = pd.read_csv(".\\sensitivityFiles\\cumInfectSeries_sc12_halfImport_r02.5_contact10c.csv",index_col = 0)
    infectTS_r25c10_full   = pd.read_csv(".\\sensitivityFiles\\cumInfectSeries_sc12_r02.5_contact10.csv",index_col = 0)
    infectTS_r25c10_double = pd.read_csv(".\\sensitivityFiles\\cumInfectSeries_sc12_doubleImport_r02.5_contact10c.csv",index_col = 0)


    rate_r15c8   = np.array([0.5*(0.8279664+9.490872),(0.8279664+9.490872),2*(0.8279664+9.490872)])
    infect_r15c8 = np.array([infectTS_r15c8_half.iloc[-1,1],infectTS_r15c8_full.iloc[-1,1],infectTS_r15c8_double.iloc[-1,1]])
    slope_r15c8  = opt.least_squares(slopeError,1.,method = 'lm',kwargs = {'predictor':rate_r15c8,'output':infect_r15c8})
    print("Scaling Relationship for R0 = 1.5, 8 contacts/day: ",slope_r15c8['x'])


    rate_r15c10   = np.array([0.5*(1.324746+8.282943),(1.324746+8.282943),2*(1.324746+8.282943)])
    infect_r15c10 = np.array([infectTS_r15c10_half.iloc[-1,1],infectTS_r15c10_full.iloc[-1,1],infectTS_r15c10_double.iloc[-1,1]])
    slope_r15c10  = opt.least_squares(slopeError,1.,method = 'lm',kwargs = {'predictor':rate_r15c10,'output':infect_r15c10})
    print("Scaling Relationship for R0 = 1.5, 10 contacts/day: ",slope_r15c10['x'])


    rate_r25c8   = np.array([0.5*(1.379944+15.81812),(1.379944+15.81812),2*(1.379944+15.81812)])
    infect_r25c8 = np.array([infectTS_r25c8_half.iloc[-1,1],infectTS_r25c8_full.iloc[-1,1],infectTS_r25c8_double.iloc[-1,1]])
    slope_r25c8  = opt.least_squares(slopeError,1.,method = 'lm',kwargs = {'predictor':rate_r25c8,'output':infect_r25c8})
    print("Scaling Relationship for R0 = 2.5, 8 contacts/day: ",slope_r25c8['x'])


    rate_r25c10    = np.array([0.5*(2.207910+13.80490),(2.207910+13.80490),2*(2.207910+13.80490)])
    infect_r25c10  = np.array([infectTS_r25c10_half.iloc[-1,1],infectTS_r25c10_full.iloc[-1,1],infectTS_r25c10_double.iloc[-1,1]])
    slope_r25c10   = opt.least_squares(slopeError,1.,method = 'lm',kwargs = {'predictor':rate_r25c10,'output':infect_r25c10})
    print("Scaling Relationship for R0 = 2.5, 10 contacts/day: ",slope_r25c10['x'])
```
