## Supplementary material for "Monitoring and responding to emerging infectious diseases in a university setting: A case study using COVID-19": S2 Supporting Information: QUCovasimAnalyses_PLOSReady.py

```
'''
This script contains the code necessary to recreate the analyses contained in Soda, Chen, Feinn, and Hill.
Author: K. James Soda
Date  : Dec. 21, 2022
'''


import sciris as sc
import covasim as cv
import covasim.simCampus as cvsc
import covasim.interventionsCampus as cvic
from datetime import datetime
from multiprocessing import freeze_support
from pandas import DataFrame
import numpy as np
import QUDormsLegacy_20200904_208 as qud


startTime = datetime.now()

# Run options
scenario =  "R0_1.5_Contacts_8"
writeInfectionToFile = True
seriesFilePath    = ".\\series.csv"
diagnosedFilePath = ".\\diagnosed.csv"
testsFilePath     = ".\\tests.csv"


if scenario == "R0_1.5_Contacts_8":
    simGroup = "new8ContactGroup"
    residGeneratorAssure = cvic.FloorTargetedPools(0.075,assureCoverage = True,skipQuarantine = True,skipDiagnosed = True,collapse = True,lock = False)
    residGeneratorNotAssure = cvic.FloorTargetedPools(0.075,assureCoverage = False,skipQuarantine = True,skipDiagnosed = True,collapse = True,lock = False)
    nonResidGenerator = cvic.RandomNonResidentSample(0.125)


    dorm_set = "nq_2020_07_24Dedense"
    numberRuns = 1000
    currentNonRes = 7425 - 3636
    nonResImportRate = 9.490872/7
    pars = sc.objdict(
        beta         = 0.023166,
        pop_infected = 0,       # Number of initial infections
        n_imports    = 0.8279664/7,
        rand_seed    = 3,        # Random seed
        pop_type     = 'campus',
        start_day = '2020-08-31',
        end_day = '2020-11-24'
    )
    pars.interventions = [cvic.symptomQuarantine(symp_prob= 0.5, #This indicates the expected time to find a symptomatic individual is one day
                                                ili_prev=None,
                                                test_sensitivity= 0.95, #Based on a nasopharyngeal swab (Hanson et al. 2020)
                                                test_delay=1, 
                                                start_day=0, 
                                                end_day=None),
                            cv.contact_tracing(trace_probs = {'r':1.,'b':0.75,'f':0.75,'c':0.75}, 
                                                trace_time = 2, 
                                                start_day  = 0, 
                                                end_day    = None, 
                                                presumptive=False),
                            cvic.TestScheduler(sampleGenerator  = residGeneratorAssure.create,
                                               schedule         = 7,
                                               start_date       = 1,
                                               end_date         = None,
                                               test_sensitivity = 0.95,
                                               test_delay       = 1, 
                                               loss_prob        = 0., 
                                               end_quarantine   = False),
                            cvic.TestScheduler(sampleGenerator  = residGeneratorNotAssure.create,
                                               schedule         = 7,
                                               start_date       = 2,
                                               end_date         = None,
                                               test_sensitivity = 0.95,
                                               test_delay       = 1, 
                                               loss_prob        = 0., 
                                               end_quarantine   = False),
                            cvic.TestScheduler(sampleGenerator  = nonResidGenerator.create,
                                               schedule         = 7,
                                               start_date       = 1,
                                               end_date         = None,
                                               test_sensitivity = 0.95,
                                               test_delay       = 1, 
                                               loss_prob        = 0., 
                                               end_quarantine   = False),
                            cvic.TestScheduler(sampleGenerator  = nonResidGenerator.create,
                                               schedule         = 7,
                                               start_date       = 2,
                                               end_date         = None,
                                               test_sensitivity = 0.95,
                                               test_delay       = 1, 
                                               loss_prob        = 0., 
                                               end_quarantine   = False)]
elif scenario == "R0_1.5_Contacts_10":
    simGroup = "BhartiUpperBound"
    residGeneratorAssure = cvic.FloorTargetedPools(0.075,assureCoverage = True,skipQuarantine = True,skipDiagnosed = True,collapse = True,lock = False)
    residGeneratorNotAssure = cvic.FloorTargetedPools(0.075,assureCoverage = False,skipQuarantine = True,skipDiagnosed = True,collapse = True,lock = False)
    nonResidGenerator = cvic.RandomNonResidentSample(0.125)


    dorm_set = "nq_2020_07_24Dedense"
    numberRuns = 1000
    currentNonRes = 7425 - 3636
    nonResImportRate = 8.282943/7
    pars = sc.objdict(
        beta         = 0.0185328,
        pop_infected = 0,       # Number of initial infections
        n_imports    = 1.324746/7,
        rand_seed    = 3,        # Random seed
        pop_type     = 'campus',
        start_day = '2020-08-31',
        end_day = '2020-11-24'
    )
    pars.interventions = [cvic.symptomQuarantine(symp_prob= 0.5, #This indicates the expected time to find a symptomatic individual is one day
                                                ili_prev=None,
                                                test_sensitivity= 0.95, #Based on a nasopharyngeal swab (Hanson et al. 2020)
                                                test_delay=1, 
                                                start_day=0, 
                                                end_day=None),
                            cv.contact_tracing(trace_probs = {'r':1.,'b':0.75,'f':0.75,'c':0.75}, 
                                                trace_time = 2, 
                                                start_day  = 0, 
                                                end_day    = None, 
                                                presumptive=False),
                            cvic.TestScheduler(sampleGenerator  = residGeneratorAssure.create,
                                               schedule         = 7,
                                               start_date       = 1,
                                               end_date         = None,
                                               test_sensitivity = 0.95,
                                               test_delay       = 1, 
                                               loss_prob        = 0., 
                                               end_quarantine   = False),
                            cvic.TestScheduler(sampleGenerator  = residGeneratorNotAssure.create,
                                               schedule         = 7,
                                               start_date       = 2,
                                               end_date         = None,
                                               test_sensitivity = 0.95,
                                               test_delay       = 1, 
                                               loss_prob        = 0., 
                                               end_quarantine   = False),
                            cvic.TestScheduler(sampleGenerator  = nonResidGenerator.create,
                                               schedule         = 7,
                                               start_date       = 1,
                                               end_date         = None,
                                               test_sensitivity = 0.95,
                                               test_delay       = 1, 
                                               loss_prob        = 0., 
                                               end_quarantine   = False),
                            cvic.TestScheduler(sampleGenerator  = nonResidGenerator.create,
                                               schedule         = 7,
                                               start_date       = 2,
                                               end_date         = None,
                                               test_sensitivity = 0.95,
                                               test_delay       = 1, 
                                               loss_prob        = 0., 
                                               end_quarantine   = False)]
elif scenario == "R0_2.5_Contacts_8":
    simGroup = "new8ContactGroup"
    residGeneratorAssure = cvic.FloorTargetedPools(0.075,assureCoverage = True,skipQuarantine = True,skipDiagnosed = True,collapse = True,lock = False)
    residGeneratorNotAssure = cvic.FloorTargetedPools(0.075,assureCoverage = False,skipQuarantine = True,skipDiagnosed = True,collapse = True,lock = False)
    nonResidGenerator = cvic.RandomNonResidentSample(0.125)


    dorm_set = "nq_2020_07_24Dedense"
    numberRuns = 1000
    currentNonRes = 7425 - 3636
    nonResImportRate = 15.81812/7
    pars = sc.objdict(
        beta         = 0.03861,
        pop_infected = 0,       # Number of initial infections
        n_imports    = 1.379944/7,
        rand_seed    = 3,        # Random seed
        pop_type     = 'campus',
        start_day = '2020-08-31',
        end_day = '2020-11-24'
    )
    pars.interventions = [cvic.symptomQuarantine(symp_prob= 0.5, #This indicates the expected time to find a symptomatic individual is one day
                                                ili_prev=None,
                                                test_sensitivity= 0.95, #Based on a nasopharyngeal swab (Hanson et al. 2020)
                                                test_delay=1, 
                                                start_day=0, 
                                                end_day=None),
                            cv.contact_tracing(trace_probs = {'r':1.,'b':0.75,'f':0.75,'c':0.75}, 
                                                trace_time = 2, 
                                                start_day  = 0, 
                                                end_day    = None, 
                                                presumptive=False),
                            cvic.TestScheduler(sampleGenerator  = residGeneratorAssure.create,
                                               schedule         = 7,
                                               start_date       = 1,
                                               end_date         = None,
                                               test_sensitivity = 0.95,
                                               test_delay       = 1, 
                                               loss_prob        = 0., 
                                               end_quarantine   = False),
                            cvic.TestScheduler(sampleGenerator  = residGeneratorNotAssure.create,
                                               schedule         = 7,
                                               start_date       = 2,
                                               end_date         = None,
                                               test_sensitivity = 0.95,
                                               test_delay       = 1, 
                                               loss_prob        = 0., 
                                               end_quarantine   = False),
                            cvic.TestScheduler(sampleGenerator  = nonResidGenerator.create,
                                               schedule         = 7,
                                               start_date       = 1,
                                               end_date         = None,
                                               test_sensitivity = 0.95,
                                               test_delay       = 1, 
                                               loss_prob        = 0., 
                                               end_quarantine   = False),
                            cvic.TestScheduler(sampleGenerator  = nonResidGenerator.create,
                                               schedule         = 7,
                                               start_date       = 2,
                                               end_date         = None,
                                               test_sensitivity = 0.95,
                                               test_delay       = 1, 
                                               loss_prob        = 0., 
                                               end_quarantine   = False)]
elif scenario == "R0_2.5_Contacts_10":
    simGroup = "BhartiUpperBound"
    residGeneratorAssure = cvic.FloorTargetedPools(0.075,assureCoverage = True,skipQuarantine = True,skipDiagnosed = True,collapse = True,lock = False)
    residGeneratorNotAssure = cvic.FloorTargetedPools(0.075,assureCoverage = False,skipQuarantine = True,skipDiagnosed = True,collapse = True,lock = False)
    nonResidGenerator = cvic.RandomNonResidentSample(0.125)


    dorm_set = "nq_2020_07_24Dedense"
    numberRuns = 1000
    currentNonRes = 7425 - 3636
    nonResImportRate = 13.80490/7
    pars = sc.objdict(
        beta         = 0.030888,
        pop_infected = 0,       # Number of initial infections
        n_imports    = 2.207910/7,
        rand_seed    = 3,        # Random seed
        pop_type     = 'campus',
        start_day = '2020-08-31',
        end_day = '2020-11-24'
    )
    pars.interventions = [cvic.symptomQuarantine(symp_prob= 0.5, #This indicates the expected time to find a symptomatic individual is one day
                                                ili_prev=None,
                                                test_sensitivity= 0.95, #Based on a nasopharyngeal swab (Hanson et al. 2020)
                                                test_delay=1, 
                                                start_day=0, 
                                                end_day=None),
                            cv.contact_tracing(trace_probs = {'r':1.,'b':0.75,'f':0.75,'c':0.75}, 
                                                trace_time = 2, 
                                                start_day  = 0, 
                                                end_day    = None, 
                                                presumptive=False),
                            cvic.TestScheduler(sampleGenerator  = residGeneratorAssure.create,
                                               schedule         = 7,
                                               start_date       = 1,
                                               end_date         = None,
                                               test_sensitivity = 0.95,
                                               test_delay       = 1, 
                                               loss_prob        = 0., 
                                               end_quarantine   = False),
                            cvic.TestScheduler(sampleGenerator  = residGeneratorNotAssure.create,
                                               schedule         = 7,
                                               start_date       = 2,
                                               end_date         = None,
                                               test_sensitivity = 0.95,
                                               test_delay       = 1, 
                                               loss_prob        = 0., 
                                               end_quarantine   = False),
                            cvic.TestScheduler(sampleGenerator  = nonResidGenerator.create,
                                               schedule         = 7,
                                               start_date       = 1,
                                               end_date         = None,
                                               test_sensitivity = 0.95,
                                               test_delay       = 1, 
                                               loss_prob        = 0., 
                                               end_quarantine   = False),
                            cvic.TestScheduler(sampleGenerator  = nonResidGenerator.create,
                                               schedule         = 7,
                                               start_date       = 2,
                                               end_date         = None,
                                               test_sensitivity = 0.95,
                                               test_delay       = 1, 
                                               loss_prob        = 0., 
                                               end_quarantine   = False)]

elif scenario == "R0_1.5_Contacts_8_HalfImport":
    simGroup = "new8ContactGroup"
    residGeneratorAssure = cvic.FloorTargetedPools(0.075,assureCoverage = True,skipQuarantine = True,skipDiagnosed = True,collapse = True,lock = False)
    residGeneratorNotAssure = cvic.FloorTargetedPools(0.075,assureCoverage = False,skipQuarantine = True,skipDiagnosed = True,collapse = True,lock = False)
    nonResidGenerator = cvic.RandomNonResidentSample(0.125)


    dorm_set = "nq_2020_07_24Dedense"
    numberRuns = 1000
    currentNonRes = 7425 - 3636
    nonResImportRate = 0.5*9.490872/7
    pars = sc.objdict(
        beta         = 0.023166,
        pop_infected = 0,       # Number of initial infections
        n_imports    = 0.5*0.8279664/7,
        rand_seed    = 3,        # Random seed
        pop_type     = 'campus',
        start_day = '2020-08-31',
        end_day = '2020-11-24'
    )
    pars.interventions = [cvic.symptomQuarantine(symp_prob= 0.5, #This indicates the expected time to find a symptomatic individual is one day
                                                ili_prev=None,
                                                test_sensitivity= 0.95, #Based on a nasopharyngeal swab (Hanson et al. 2020)
                                                test_delay=1, 
                                                start_day=0, 
                                                end_day=None),
                            cv.contact_tracing(trace_probs = {'r':1.,'b':0.75,'f':0.75,'c':0.75}, 
                                                trace_time = 2, 
                                                start_day  = 0, 
                                                end_day    = None, 
                                                presumptive=False),
                            cvic.TestScheduler(sampleGenerator  = residGeneratorAssure.create,
                                               schedule         = 7,
                                               start_date       = 1,
                                               end_date         = None,
                                               test_sensitivity = 0.95,
                                               test_delay       = 1, 
                                               loss_prob        = 0., 
                                               end_quarantine   = False),
                            cvic.TestScheduler(sampleGenerator  = residGeneratorNotAssure.create,
                                               schedule         = 7,
                                               start_date       = 2,
                                               end_date         = None,
                                               test_sensitivity = 0.95,
                                               test_delay       = 1, 
                                               loss_prob        = 0., 
                                               end_quarantine   = False),
                            cvic.TestScheduler(sampleGenerator  = nonResidGenerator.create,
                                               schedule         = 7,
                                               start_date       = 1,
                                               end_date         = None,
                                               test_sensitivity = 0.95,
                                               test_delay       = 1, 
                                               loss_prob        = 0., 
                                               end_quarantine   = False),
                            cvic.TestScheduler(sampleGenerator  = nonResidGenerator.create,
                                               schedule         = 7,
                                               start_date       = 2,
                                               end_date         = None,
                                               test_sensitivity = 0.95,
                                               test_delay       = 1, 
                                               loss_prob        = 0., 
                                               end_quarantine   = False)]

elif scenario == "R0_1.5_Contacts_8_DoubleImport":
    simGroup = "new8ContactGroup"
    residGeneratorAssure = cvic.FloorTargetedPools(0.075,assureCoverage = True,skipQuarantine = True,skipDiagnosed = True,collapse = True,lock = False)
    residGeneratorNotAssure = cvic.FloorTargetedPools(0.075,assureCoverage = False,skipQuarantine = True,skipDiagnosed = True,collapse = True,lock = False)
    nonResidGenerator = cvic.RandomNonResidentSample(0.125)


    dorm_set = "nq_2020_07_24Dedense"
    numberRuns = 1000
    currentNonRes = 7425 - 3636
    nonResImportRate = 2*9.490872/7
    pars = sc.objdict(
        beta         = 0.023166,
        pop_infected = 0,       # Number of initial infections
        n_imports    = 2*0.8279664/7,
        rand_seed    = 3,        # Random seed
        pop_type     = 'campus',
        start_day = '2020-08-31',
        end_day = '2020-11-24'
    )
    pars.interventions = [cvic.symptomQuarantine(symp_prob= 0.5, #This indicates the expected time to find a symptomatic individual is one day
                                                ili_prev=None,
                                                test_sensitivity= 0.95, #Based on a nasopharyngeal swab (Hanson et al. 2020)
                                                test_delay=1, 
                                                start_day=0, 
                                                end_day=None),
                            cv.contact_tracing(trace_probs = {'r':1.,'b':0.75,'f':0.75,'c':0.75}, 
                                                trace_time = 2, 
                                                start_day  = 0, 
                                                end_day    = None, 
                                                presumptive=False),
                            cvic.TestScheduler(sampleGenerator  = residGeneratorAssure.create,
                                               schedule         = 7,
                                               start_date       = 1,
                                               end_date         = None,
                                               test_sensitivity = 0.95,
                                               test_delay       = 1, 
                                               loss_prob        = 0., 
                                               end_quarantine   = False),
                            cvic.TestScheduler(sampleGenerator  = residGeneratorNotAssure.create,
                                               schedule         = 7,
                                               start_date       = 2,
                                               end_date         = None,
                                               test_sensitivity = 0.95,
                                               test_delay       = 1, 
                                               loss_prob        = 0., 
                                               end_quarantine   = False),
                            cvic.TestScheduler(sampleGenerator  = nonResidGenerator.create,
                                               schedule         = 7,
                                               start_date       = 1,
                                               end_date         = None,
                                               test_sensitivity = 0.95,
                                               test_delay       = 1, 
                                               loss_prob        = 0., 
                                               end_quarantine   = False),
                            cvic.TestScheduler(sampleGenerator  = nonResidGenerator.create,
                                               schedule         = 7,
                                               start_date       = 2,
                                               end_date         = None,
                                               test_sensitivity = 0.95,
                                               test_delay       = 1, 
                                               loss_prob        = 0., 
                                               end_quarantine   = False)]

elif scenario == "R0_1.5_Contacts_10_HalfImport":
    simGroup = "BhartiUpperBound"
    residGeneratorAssure = cvic.FloorTargetedPools(0.075,assureCoverage = True,skipQuarantine = True,skipDiagnosed = True,collapse = True,lock = False)
    residGeneratorNotAssure = cvic.FloorTargetedPools(0.075,assureCoverage = False,skipQuarantine = True,skipDiagnosed = True,collapse = True,lock = False)

    nonResidGenerator = cvic.RandomNonResidentSample(0.125)


    dorm_set = "nq_2020_07_24Dedense"
    numberRuns = 1000
    currentNonRes = 7425 - 3636
    nonResImportRate = 0.5*8.282943/7
    pars = sc.objdict(
        beta         = 0.0185328,
        pop_infected = 0,       # Number of initial infections
        n_imports    = 0.5*1.324746/7,
        rand_seed    = 3,        # Random seed
        pop_type     = 'campus',
        start_day = '2020-08-31',
        end_day = '2020-11-24'
    )
    pars.interventions = [cvic.symptomQuarantine(symp_prob= 0.5, #This indicates the expected time to find a symptomatic individual is one day
                                                ili_prev=None,
                                                test_sensitivity= 0.95, #Based on a nasopharyngeal swab (Hanson et al. 2020)
                                                test_delay=1, 
                                                start_day=0, 
                                                end_day=None),
                            cv.contact_tracing(trace_probs = {'r':1.,'b':0.75,'f':0.75,'c':0.75}, 
                                                trace_time = 2, 
                                                start_day  = 0, 
                                                end_day    = None, 
                                                presumptive=False),
                            cvic.TestScheduler(sampleGenerator  = residGeneratorAssure.create,
                                               schedule         = 7,
                                               start_date       = 1,
                                               end_date         = None,
                                               test_sensitivity = 0.95,
                                               test_delay       = 1, 
                                               loss_prob        = 0., 
                                               end_quarantine   = False),
                            cvic.TestScheduler(sampleGenerator  = residGeneratorNotAssure.create,
                                               schedule         = 7,
                                               start_date       = 2,
                                               end_date         = None,
                                               test_sensitivity = 0.95,
                                               test_delay       = 1, 
                                               loss_prob        = 0., 
                                               end_quarantine   = False),
                            cvic.TestScheduler(sampleGenerator  = nonResidGenerator.create,
                                               schedule         = 7,
                                               start_date       = 1,
                                               end_date         = None,
                                               test_sensitivity = 0.95,
                                               test_delay       = 1, 
                                               loss_prob        = 0., 
                                               end_quarantine   = False),
                            cvic.TestScheduler(sampleGenerator  = nonResidGenerator.create,
                                               schedule         = 7,
                                               start_date       = 2,
                                               end_date         = None,
                                               test_sensitivity = 0.95,
                                               test_delay       = 1, 
                                               loss_prob        = 0., 
                                               end_quarantine   = False)]

elif scenario == "R0_1.5_Contacts_10_DoubleImport":
    simGroup = "BhartiUpperBound"
    residGeneratorAssure = cvic.FloorTargetedPools(0.075,assureCoverage = True,skipQuarantine = True,skipDiagnosed = True,collapse = True,lock = False)
    residGeneratorNotAssure = cvic.FloorTargetedPools(0.075,assureCoverage = False,skipQuarantine = True,skipDiagnosed = True,collapse = True,lock = False)
    nonResidGenerator = cvic.RandomNonResidentSample(0.125)


    dorm_set = "nq_2020_07_24Dedense"
    numberRuns = 1000
    currentNonRes = 7425 - 3636
    nonResImportRate = 2*8.282943/7
    pars = sc.objdict(
        beta         = 0.0185328,
        pop_infected = 0,       # Number of initial infections
        n_imports    = 2*1.324746/7,
        rand_seed    = 3,        # Random seed
        pop_type     = 'campus',
        start_day = '2020-08-31',
        end_day = '2020-11-24'
    )
    pars.interventions = [cvic.symptomQuarantine(symp_prob= 0.5, #This indicates the expected time to find a symptomatic individual is one day
                                                ili_prev=None,
                                                test_sensitivity= 0.95, #Based on a nasopharyngeal swab (Hanson et al. 2020)
                                                test_delay=1, 
                                                start_day=0, 
                                                end_day=None),
                            cv.contact_tracing(trace_probs = {'r':1.,'b':0.75,'f':0.75,'c':0.75}, 
                                                trace_time = 2, 
                                                start_day  = 0, 
                                                end_day    = None, 
                                                presumptive=False),
                            cvic.TestScheduler(sampleGenerator  = residGeneratorAssure.create,
                                               schedule         = 7,
                                               start_date       = 1,
                                               end_date         = None,
                                               test_sensitivity = 0.95,
                                               test_delay       = 1, 
                                               loss_prob        = 0., 
                                               end_quarantine   = False),
                            cvic.TestScheduler(sampleGenerator  = residGeneratorNotAssure.create,
                                               schedule         = 7,
                                               start_date       = 2,
                                               end_date         = None,
                                               test_sensitivity = 0.95,
                                               test_delay       = 1, 
                                               loss_prob        = 0., 
                                               end_quarantine   = False),
                            cvic.TestScheduler(sampleGenerator  = nonResidGenerator.create,
                                               schedule         = 7,
                                               start_date       = 1,
                                               end_date         = None,
                                               test_sensitivity = 0.95,
                                               test_delay       = 1, 
                                               loss_prob        = 0., 
                                               end_quarantine   = False),
                            cvic.TestScheduler(sampleGenerator  = nonResidGenerator.create,
                                               schedule         = 7,
                                               start_date       = 2,
                                               end_date         = None,
                                               test_sensitivity = 0.95,
                                               test_delay       = 1, 
                                               loss_prob        = 0., 
                                               end_quarantine   = False)]

elif scenario == "R0_2.5_Contacts_8_HalfImport":
    simGroup = "new8ContactGroup"
    residGeneratorAssure = cvic.FloorTargetedPools(0.075,assureCoverage = True,skipQuarantine = True,skipDiagnosed = True,collapse = True,lock = False)
    residGeneratorNotAssure = cvic.FloorTargetedPools(0.075,assureCoverage = False,skipQuarantine = True,skipDiagnosed = True,collapse = True,lock = False)
    nonResidGenerator = cvic.RandomNonResidentSample(0.125)


    dorm_set = "nq_2020_07_24Dedense"
    numberRuns = 1000
    currentNonRes = 7425 - 3636
    nonResImportRate = 0.5*15.81812/7
    pars = sc.objdict(
        beta         = 0.03861,
        pop_infected = 0,       # Number of initial infections
        n_imports    = 0.5*1.379944/7,
        rand_seed    = 3,        # Random seed
        pop_type     = 'campus',
        start_day = '2020-08-31',
        end_day = '2020-11-24'
    )
    pars.interventions = [cvic.symptomQuarantine(symp_prob= 0.5, #This indicates the expected time to find a symptomatic individual is one day
                                                ili_prev=None,
                                                test_sensitivity= 0.95, #Based on a nasopharyngeal swab (Hanson et al. 2020)
                                                test_delay=1, 
                                                start_day=0, 
                                                end_day=None),
                            cv.contact_tracing(trace_probs = {'r':1.,'b':0.75,'f':0.75,'c':0.75}, 
                                                trace_time = 2, 
                                                start_day  = 0, 
                                                end_day    = None, 
                                                presumptive=False),
                            cvic.TestScheduler(sampleGenerator  = residGeneratorAssure.create,
                                               schedule         = 7,
                                               start_date       = 1,
                                               end_date         = None,
                                               test_sensitivity = 0.95,
                                               test_delay       = 1, 
                                               loss_prob        = 0., 
                                               end_quarantine   = False),
                            cvic.TestScheduler(sampleGenerator  = residGeneratorNotAssure.create,
                                               schedule         = 7,
                                               start_date       = 2,
                                               end_date         = None,
                                               test_sensitivity = 0.95,
                                               test_delay       = 1, 
                                               loss_prob        = 0., 
                                               end_quarantine   = False),
                            cvic.TestScheduler(sampleGenerator  = nonResidGenerator.create,
                                               schedule         = 7,
                                               start_date       = 1,
                                               end_date         = None,
                                               test_sensitivity = 0.95,
                                               test_delay       = 1, 
                                               loss_prob        = 0., 
                                               end_quarantine   = False),
                            cvic.TestScheduler(sampleGenerator  = nonResidGenerator.create,
                                               schedule         = 7,
                                               start_date       = 2,
                                               end_date         = None,
                                               test_sensitivity = 0.95,
                                               test_delay       = 1, 
                                               loss_prob        = 0., 
                                               end_quarantine   = False)]

elif scenario == "R0_2.5_Contacts_8_DoubleImport":
    simGroup = "new8ContactGroup"
    residGeneratorAssure = cvic.FloorTargetedPools(0.075,assureCoverage = True,skipQuarantine = True,skipDiagnosed = True,collapse = True,lock = False)
    residGeneratorNotAssure = cvic.FloorTargetedPools(0.075,assureCoverage = False,skipQuarantine = True,skipDiagnosed = True,collapse = True,lock = False)
    nonResidGenerator = cvic.RandomNonResidentSample(0.125)


    dorm_set = "nq_2020_07_24Dedense"
    numberRuns = 1000
    currentNonRes = 7425 - 3636
    nonResImportRate = 2*15.81812/7
    pars = sc.objdict(
        beta         = 0.03861,
        pop_infected = 0,       # Number of initial infections
        n_imports    = 2*1.379944/7,
        rand_seed    = 3,        # Random seed
        pop_type     = 'campus',
        start_day = '2020-08-31',
        end_day = '2020-11-24'
    )
    pars.interventions = [cvic.symptomQuarantine(symp_prob= 0.5, #This indicates the expected time to find a symptomatic individual is one day
                                                ili_prev=None,
                                                test_sensitivity= 0.95, #Based on a nasopharyngeal swab (Hanson et al. 2020)
                                                test_delay=1, 
                                                start_day=0, 
                                                end_day=None),
                            cv.contact_tracing(trace_probs = {'r':1.,'b':0.75,'f':0.75,'c':0.75}, 
                                                trace_time = 2, 
                                                start_day  = 0, 
                                                end_day    = None, 
                                                presumptive=False),
                            cvic.TestScheduler(sampleGenerator  = residGeneratorAssure.create,
                                               schedule         = 7,
                                               start_date       = 1,
                                               end_date         = None,
                                               test_sensitivity = 0.95,
                                               test_delay       = 1, 
                                               loss_prob        = 0., 
                                               end_quarantine   = False),
                            cvic.TestScheduler(sampleGenerator  = residGeneratorNotAssure.create,
                                               schedule         = 7,
                                               start_date       = 2,
                                               end_date         = None,
                                               test_sensitivity = 0.95,
                                               test_delay       = 1, 
                                               loss_prob        = 0., 
                                               end_quarantine   = False),
                            cvic.TestScheduler(sampleGenerator  = nonResidGenerator.create,
                                               schedule         = 7,
                                               start_date       = 1,
                                               end_date         = None,
                                               test_sensitivity = 0.95,
                                               test_delay       = 1, 
                                               loss_prob        = 0., 
                                               end_quarantine   = False),
                            cvic.TestScheduler(sampleGenerator  = nonResidGenerator.create,
                                               schedule         = 7,
                                               start_date       = 2,
                                               end_date         = None,
                                               test_sensitivity = 0.95,
                                               test_delay       = 1, 
                                               loss_prob        = 0., 
                                               end_quarantine   = False)]

elif scenario == "R0_2.5_Contacts_10_HalfImport":
    simGroup = "BhartiUpperBound"
    residGeneratorAssure = cvic.FloorTargetedPools(0.075,assureCoverage = True,skipQuarantine = True,skipDiagnosed = True,collapse = True,lock = False)
    residGeneratorNotAssure = cvic.FloorTargetedPools(0.075,assureCoverage = False,skipQuarantine = True,skipDiagnosed = True,collapse = True,lock = False)
    nonResidGenerator = cvic.RandomNonResidentSample(0.125)


    dorm_set = "nq_2020_07_24Dedense"
    numberRuns = 1000
    currentNonRes = 7425 - 3636
    nonResImportRate = 0.5*13.80490/7
    pars = sc.objdict(
        beta         = 0.030888,
        pop_infected = 0,       # Number of initial infections
        n_imports    = 0.5*2.207910/7,
        rand_seed    = 3,        # Random seed
        pop_type     = 'campus',
        start_day = '2020-08-31',
        end_day = '2020-11-24'
    )
    pars.interventions = [cvic.symptomQuarantine(symp_prob= 0.5, #This indicates the expected time to find a symptomatic individual is one day
                                                ili_prev=None,
                                                test_sensitivity= 0.95, #Based on a nasopharyngeal swab (Hanson et al. 2020)
                                                test_delay=1, 
                                                start_day=0, 
                                                end_day=None),
                            cv.contact_tracing(trace_probs = {'r':1.,'b':0.75,'f':0.75,'c':0.75}, 
                                                trace_time = 2, 
                                                start_day  = 0, 
                                                end_day    = None, 
                                                presumptive=False),
                            cvic.TestScheduler(sampleGenerator  = residGeneratorAssure.create,
                                               schedule         = 7,
                                               start_date       = 1,
                                               end_date         = None,
                                               test_sensitivity = 0.95,
                                               test_delay       = 1, 
                                               loss_prob        = 0., 
                                               end_quarantine   = False),
                            cvic.TestScheduler(sampleGenerator  = residGeneratorNotAssure.create,
                                               schedule         = 7,
                                               start_date       = 2,
                                               end_date         = None,
                                               test_sensitivity = 0.95,
                                               test_delay       = 1, 
                                               loss_prob        = 0., 
                                               end_quarantine   = False),
                            cvic.TestScheduler(sampleGenerator  = nonResidGenerator.create,
                                               schedule         = 7,
                                               start_date       = 1,
                                               end_date         = None,
                                               test_sensitivity = 0.95,
                                               test_delay       = 1, 
                                               loss_prob        = 0., 
                                               end_quarantine   = False),
                            cvic.TestScheduler(sampleGenerator  = nonResidGenerator.create,
                                               schedule         = 7,
                                               start_date       = 2,
                                               end_date         = None,
                                               test_sensitivity = 0.95,
                                               test_delay       = 1, 
                                               loss_prob        = 0., 
                                               end_quarantine   = False)]

elif scenario == "R0_2.5_Contacts_10_DoubleImport":
    simGroup = "BhartiUpperBound"
    residGeneratorAssure = cvic.FloorTargetedPools(0.075,assureCoverage = True,skipQuarantine = True,skipDiagnosed = True,collapse = True,lock = False)
    residGeneratorNotAssure = cvic.FloorTargetedPools(0.075,assureCoverage = False,skipQuarantine = True,skipDiagnosed = True,collapse = True,lock = False)
    nonResidGenerator = cvic.RandomNonResidentSample(0.125)


    dorm_set = "nq_2020_07_24Dedense"
    numberRuns = 1000
    currentNonRes = 7425 - 3636
    nonResImportRate = 2*13.80490/7
    pars = sc.objdict(
        beta         = 0.030888,
        pop_infected = 0,       # Number of initial infections
        n_imports    = 2*2.207910/7,
        rand_seed    = 3,        # Random seed
        pop_type     = 'campus',
        start_day = '2020-08-31',
        end_day = '2020-11-24'
    )
    pars.interventions = [cvic.symptomQuarantine(symp_prob= 0.5, #This indicates the expected time to find a symptomatic individual is one day
                                                ili_prev=None,
                                                test_sensitivity= 0.95, #Based on a nasopharyngeal swab (Hanson et al. 2020)
                                                test_delay=1, 
                                                start_day=0, 
                                                end_day=None),
                            cv.contact_tracing(trace_probs = {'r':1.,'b':0.75,'f':0.75,'c':0.75}, 
                                                trace_time = 2, 
                                                start_day  = 0, 
                                                end_day    = None, 
                                                presumptive=False),
                            cvic.TestScheduler(sampleGenerator  = residGeneratorAssure.create,
                                               schedule         = 7,
                                               start_date       = 1,
                                               end_date         = None,
                                               test_sensitivity = 0.95,
                                               test_delay       = 1, 
                                               loss_prob        = 0., 
                                               end_quarantine   = False),
                            cvic.TestScheduler(sampleGenerator  = residGeneratorNotAssure.create,
                                               schedule         = 7,
                                               start_date       = 2,
                                               end_date         = None,
                                               test_sensitivity = 0.95,
                                               test_delay       = 1, 
                                               loss_prob        = 0., 
                                               end_quarantine   = False),
                            cvic.TestScheduler(sampleGenerator  = nonResidGenerator.create,
                                               schedule         = 7,
                                               start_date       = 1,
                                               end_date         = None,
                                               test_sensitivity = 0.95,
                                               test_delay       = 1, 
                                               loss_prob        = 0., 
                                               end_quarantine   = False),
                            cvic.TestScheduler(sampleGenerator  = nonResidGenerator.create,
                                               schedule         = 7,
                                               start_date       = 2,
                                               end_date         = None,
                                               test_sensitivity = 0.95,
                                               test_delay       = 1, 
                                               loss_prob        = 0., 
                                               end_quarantine   = False)]

elif scenario == "R0_1.5_Contacts_8_SRS":
    class RandomResidentSample_SetTests:
        def __init__(self,numberTests,lock = False):
            self.numberTests  = numberTests
            self.lock         = lock
            self.sample       = np.array([])

        def create(self,sim):
            if not (self.lock and len(self.sample) > 0):
                resPop  = sim.dorm_offsets[-1]
                self.sample   = np.random.choice(resPop,self.numberTests,replace = False)
            assert all(self.sample < resPop)
            return self.sample


    simGroup = "new8ContactGroup"
    residGenerator_T = RandomResidentSample_SetTests(355)
    residGenerator_W = RandomResidentSample_SetTests(225)
    nonResidGenerator = cvic.RandomNonResidentSample(0.125)


    dorm_set = "nq_2020_07_24Dedense"
    numberRuns = 1000
    currentNonRes = 7425 - 3636
    nonResImportRate = 9.490872/7
    pars = sc.objdict(
        beta         = 0.023166,
        pop_infected = 0,       # Number of initial infections
        n_imports    = 0.8279664/7,
        rand_seed    = 3,        # Random seed
        pop_type     = 'campus',
        start_day = '2020-08-31',
        end_day = '2020-11-24'
    )
    pars.interventions = [cvic.symptomQuarantine(symp_prob= 0.5, #This indicates the expected time to find a symptomatic individual is one day
                                                ili_prev=None,
                                                test_sensitivity= 0.95, #Based on a nasopharyngeal swab (Hanson et al. 2020)
                                                test_delay=1, 
                                                start_day=0, 
                                                end_day=None),
                            cv.contact_tracing(trace_probs = {'r':1.,'b':0.75,'f':0.75,'c':0.75}, 
                                                trace_time = 2, 
                                                start_day  = 0, 
                                                end_day    = None, 
                                                presumptive=False),
                            cvic.TestScheduler(sampleGenerator  = residGenerator_T.create,
                                               schedule         = 7,
                                               start_date       = 1,
                                               end_date         = None,
                                               test_sensitivity = 0.95,
                                               test_delay       = 1, 
                                               loss_prob        = 0., 
                                               end_quarantine   = False),
                            cvic.TestScheduler(sampleGenerator  = residGenerator_W.create,
                                               schedule         = 7,
                                               start_date       = 2,
                                               end_date         = None,
                                               test_sensitivity = 0.95,
                                               test_delay       = 1, 
                                               loss_prob        = 0., 
                                               end_quarantine   = False),
                            cvic.TestScheduler(sampleGenerator  = nonResidGenerator.create,
                                               schedule         = 7,
                                               start_date       = 1,
                                               end_date         = None,
                                               test_sensitivity = 0.95,
                                               test_delay       = 1, 
                                               loss_prob        = 0., 
                                               end_quarantine   = False),
                            cvic.TestScheduler(sampleGenerator  = nonResidGenerator.create,
                                               schedule         = 7,
                                               start_date       = 2,
                                               end_date         = None,
                                               test_sensitivity = 0.95,
                                               test_delay       = 1, 
                                               loss_prob        = 0., 
                                               end_quarantine   = False)]

elif scenario == "R0_1.5_Contacts_8_100percent":
    simGroup = "new8ContactGroup"
    np.random.seed(219222689)
    tuesdayPool   = np.random.choice(7425,3712,replace = False)
    wednesdayPool = np.array([i for i in range(7425) if not i in tuesdayPool])


    dorm_set = "nq_2020_07_24Dedense"
    numberRuns = 1000

    currentNonRes = 7425 - 3636
    nonResImportRate = 9.490872/7
    pars = sc.objdict(
        beta         = 0.023166,
        pop_infected = 0,       # Number of initial infections
        n_imports    = 0.8279664/7,
        rand_seed    = 3,        # Random seed
        pop_type     = 'campus',
        start_day = '2020-08-31',
        end_day = '2020-11-24'
    )
    pars.interventions = [cvic.symptomQuarantine(symp_prob= 0.5, #This indicates the expected time to find a symptomatic individual is one day
                                                ili_prev=None,
                                                test_sensitivity= 0.95, #Based on a nasopharyngeal swab (Hanson et al. 2020)
                                                test_delay=1, 
                                                start_day=0, 
                                                end_day=None),
                            cv.contact_tracing(trace_probs = {'r':1.,'b':0.75,'f':0.75,'c':0.75}, 
                                                trace_time = 2, 
                                                start_day  = 0, 
                                                end_day    = None, 
                                                presumptive=False),
                            cvic.TestScheduler(sampleGenerator  = tuesdayPool,
                                               schedule         = 7,
                                               start_date       = 1,
                                               end_date         = None,
                                               test_sensitivity = 0.95,
                                               test_delay       = 1, 
                                               loss_prob        = 0., 
                                               end_quarantine   = False),
                            cvic.TestScheduler(sampleGenerator  = wednesdayPool,
                                               schedule         = 7,
                                               start_date       = 2,
                                               end_date         = None,
                                               test_sensitivity = 0.95,
                                               test_delay       = 1, 
                                               loss_prob        = 0., 
                                               end_quarantine   = False)]

elif scenario == "R0_1.5_Contacts_10_SRS":
    class RandomResidentSample_SetTests:
        def __init__(self,numberTests,lock = False):
            self.numberTests  = numberTests
            self.lock         = lock
            self.sample       = np.array([])

        def create(self,sim):
            if not (self.lock and len(self.sample) > 0):
                resPop  = sim.dorm_offsets[-1]
                self.sample   = np.random.choice(resPop,self.numberTests,replace = False)
            assert all(self.sample < resPop)
            return self.sample


    simGroup = "BhartiUpperBound"
    residGenerator_T = RandomResidentSample_SetTests(355)
    residGenerator_W = RandomResidentSample_SetTests(225)
    nonResidGenerator = cvic.RandomNonResidentSample(0.125)


    dorm_set = "nq_2020_07_24Dedense"
    numberRuns = 1000
    currentNonRes = 7425 - 3636
    nonResImportRate = 8.282943/7
    pars = sc.objdict(
        beta         = 0.0185328,
        pop_infected = 0,       # Number of initial infections
        n_imports    = 1.324746/7,
        rand_seed    = 3,        # Random seed
        pop_type     = 'campus',
        start_day = '2020-08-31',
        end_day = '2020-11-24'
    )
    pars.interventions = [cvic.symptomQuarantine(symp_prob= 0.5, #This indicates the expected time to find a symptomatic individual is one day
                                                ili_prev=None,
                                                test_sensitivity= 0.95, #Based on a nasopharyngeal swab (Hanson et al. 2020)
                                                test_delay=1, 
                                                start_day=0, 
                                                end_day=None),
                            cv.contact_tracing(trace_probs = {'r':1.,'b':0.75,'f':0.75,'c':0.75}, 
                                                trace_time = 2, 
                                                start_day  = 0, 
                                                end_day    = None, 
                                                presumptive=False),
                            cvic.TestScheduler(sampleGenerator  = residGenerator_T.create,
                                               schedule         = 7,
                                               start_date       = 1,
                                               end_date         = None,
                                               test_sensitivity = 0.95,
                                               test_delay       = 1, 
                                               loss_prob        = 0., 
                                               end_quarantine   = False),
                            cvic.TestScheduler(sampleGenerator  = residGenerator_W.create,
                                               schedule         = 7,
                                               start_date       = 2,
                                               end_date         = None,
                                               test_sensitivity = 0.95,
                                               test_delay       = 1, 
                                               loss_prob        = 0., 
                                               end_quarantine   = False),
                            cvic.TestScheduler(sampleGenerator  = nonResidGenerator.create,
                                               schedule         = 7,
                                               start_date       = 1,
                                               end_date         = None,
                                               test_sensitivity = 0.95,
                                               test_delay       = 1, 
                                               loss_prob        = 0., 
                                               end_quarantine   = False),
                            cvic.TestScheduler(sampleGenerator  = nonResidGenerator.create,
                                               schedule         = 7,
                                               start_date       = 2,
                                               end_date         = None,
                                               test_sensitivity = 0.95,
                                               test_delay       = 1, 
                                               loss_prob        = 0., 
                                               end_quarantine   = False)]

elif scenario == "R0_1.5_Contacts_10_100percent":
    simGroup = "BhartiUpperBound"
    np.random.seed(219222689)
    tuesdayPool   = np.random.choice(7425,3712,replace = False)
    wednesdayPool = np.array([i for i in range(7425) if not i in tuesdayPool])


    dorm_set = "nq_2020_07_24Dedense"
    numberRuns = 1000

    currentNonRes = 7425 - 3636
    nonResImportRate = 8.282943/7
    pars = sc.objdict(
        beta         = 0.0185328,
        pop_infected = 0,       # Number of initial infections
        n_imports    = 1.324746/7,
        rand_seed    = 3,        # Random seed
        pop_type     = 'campus',
        start_day = '2020-08-31',
        end_day = '2020-11-24'
    )
    pars.interventions = [cvic.symptomQuarantine(symp_prob= 0.5, #This indicates the expected time to find a symptomatic individual is one day
                                                ili_prev=None,
                                                test_sensitivity= 0.95, #Based on a nasopharyngeal swab (Hanson et al. 2020)
                                                test_delay=1, 
                                                start_day=0, 
                                                end_day=None),
                            cv.contact_tracing(trace_probs = {'r':1.,'b':0.75,'f':0.75,'c':0.75}, 
                                                trace_time = 2, 
                                                start_day  = 0, 
                                                end_day    = None, 
                                                presumptive=False),
                            cvic.TestScheduler(sampleGenerator  = tuesdayPool,
                                               schedule         = 7,
                                               start_date       = 1,
                                               end_date         = None,
                                               test_sensitivity = 0.95,
                                               test_delay       = 1, 
                                               loss_prob        = 0., 
                                               end_quarantine   = False),
                            cvic.TestScheduler(sampleGenerator  = wednesdayPool,
                                               schedule         = 7,
                                               start_date       = 2,
                                               end_date         = None,
                                               test_sensitivity = 0.95,
                                               test_delay       = 1, 
                                               loss_prob        = 0., 
                                               end_quarantine   = False)]

elif scenario == "R0_2.5_Contacts_8_SRS":
    class RandomResidentSample_SetTests:
        def __init__(self,numberTests,lock = False):
            self.numberTests  = numberTests
            self.lock         = lock
            self.sample       = np.array([])

        def create(self,sim):
            if not (self.lock and len(self.sample) > 0):
                resPop  = sim.dorm_offsets[-1]
                self.sample   = np.random.choice(resPop,self.numberTests,replace = False)
            assert all(self.sample < resPop)
            return self.sample


    simGroup = "new8ContactGroup"
    residGenerator_T = RandomResidentSample_SetTests(355)
    residGenerator_W = RandomResidentSample_SetTests(225)
    nonResidGenerator = cvic.RandomNonResidentSample(0.125)


    dorm_set = "nq_2020_07_24Dedense"
    numberRuns = 1000
    currentNonRes = 7425 - 3636
    nonResImportRate = 15.81812/7
    pars = sc.objdict(
        beta         = 0.03861,
        pop_infected = 0,       # Number of initial infections
        n_imports    = 1.379944/7,
        rand_seed    = 3,        # Random seed
        pop_type     = 'campus',
        start_day = '2020-08-31',
        end_day = '2020-11-24'
    )
    pars.interventions = [cvic.symptomQuarantine(symp_prob= 0.5, #This indicates the expected time to find a symptomatic individual is one day
                                                ili_prev=None,
                                                test_sensitivity= 0.95, #Based on a nasopharyngeal swab (Hanson et al. 2020)
                                                test_delay=1, 
                                                start_day=0, 
                                                end_day=None),
                          cv.contact_tracing(trace_probs = {'r':1.,'b':0.75,'f':0.75,'c':0.75}, 
                                                trace_time = 2, 
                                                start_day  = 0, 
                                                end_day    = None, 
                                                presumptive=False),
                            cvic.TestScheduler(sampleGenerator  = residGenerator_T.create,
                                               schedule         = 7,
                                               start_date       = 1,
                                               end_date         = None,
                                               test_sensitivity = 0.95,
                                               test_delay       = 1, 
                                               loss_prob        = 0., 
                                               end_quarantine   = False),
                            cvic.TestScheduler(sampleGenerator  = residGenerator_W.create,
                                               schedule         = 7,
                                               start_date       = 2,
                                               end_date         = None,
                                               test_sensitivity = 0.95,
                                               test_delay       = 1, 
                                               loss_prob        = 0., 
                                               end_quarantine   = False),
                            cvic.TestScheduler(sampleGenerator  = nonResidGenerator.create,
                                               schedule         = 7,
                                               start_date       = 1,
                                               end_date         = None,
                                               test_sensitivity = 0.95,
                                               test_delay       = 1, 
                                               loss_prob        = 0., 
                                               end_quarantine   = False),
                            cvic.TestScheduler(sampleGenerator  = nonResidGenerator.create,
                                               schedule         = 7,
                                               start_date       = 2,
                                               end_date         = None,
                                               test_sensitivity = 0.95,
                                               test_delay       = 1, 
                                               loss_prob        = 0., 
                                               end_quarantine   = False)]

elif scenario == "R0_2.5_Contacts_8_100percent":
    simGroup = "new8ContactGroup"
    np.random.seed(219222689)
    tuesdayPool   = np.random.choice(7425,3712,replace = False)
    wednesdayPool = np.array([i for i in range(7425) if not i in tuesdayPool])


    dorm_set = "nq_2020_07_24Dedense"
    numberRuns = 1000

    currentNonRes = 7425 - 3636
    nonResImportRate = 15.81812/7
    pars = sc.objdict(
        beta         = 0.03861,
        pop_infected = 0,       # Number of initial infections
        n_imports    = 1.379944/7,
        rand_seed    = 3,        # Random seed
        pop_type     = 'campus',
        start_day = '2020-08-31',
        end_day = '2020-11-24'
    )
    pars.interventions = [cvic.symptomQuarantine(symp_prob= 0.5, #This indicates the expected time to find a symptomatic individual is one day
                                                ili_prev=None,
                                                test_sensitivity= 0.95, #Based on a nasopharyngeal swab (Hanson et al. 2020)
                                                test_delay=1, 
                                                start_day=0, 
                                                end_day=None),
                            cv.contact_tracing(trace_probs = {'r':1.,'b':0.75,'f':0.75,'c':0.75}, 
                                                trace_time = 2, 
                                                start_day  = 0, 
                                                end_day    = None, 
                                                presumptive=False),
                            cvic.TestScheduler(sampleGenerator  = tuesdayPool,
                                               schedule         = 7,
                                               start_date       = 1,
                                               end_date         = None,
                                               test_sensitivity = 0.95,
                                               test_delay       = 1, 
                                               loss_prob        = 0., 
                                               end_quarantine   = False),
                            cvic.TestScheduler(sampleGenerator  = wednesdayPool,
                                               schedule         = 7,
                                               start_date       = 2,
                                               end_date         = None,
                                               test_sensitivity = 0.95,
                                               test_delay       = 1, 
                                               loss_prob        = 0., 
                                               end_quarantine   = False)]

elif scenario == "R0_2.5_Contacts_10_SRS":
    class RandomResidentSample_SetTests:
        def __init__(self,numberTests,lock = False):
            self.numberTests  = numberTests
            self.lock         = lock
            self.sample       = np.array([])

        def create(self,sim):
            if not (self.lock and len(self.sample) > 0):
                resPop  = sim.dorm_offsets[-1]
                self.sample   = np.random.choice(resPop,self.numberTests,replace = False)
            assert all(self.sample < resPop)
            return self.sample


    simGroup = "BhartiUpperBound"
    residGenerator_T = RandomResidentSample_SetTests(355)
    residGenerator_W = RandomResidentSample_SetTests(225)
    nonResidGenerator = cvic.RandomNonResidentSample(0.125)


    dorm_set = "nq_2020_07_24Dedense"
    numberRuns = 1000
    currentNonRes = 7425 - 3636
    nonResImportRate = 13.80490/7
    pars = sc.objdict(
        beta         = 0.030888,
        pop_infected = 0,       # Number of initial infections
        n_imports    = 2.207910/7,
        rand_seed    = 3,        # Random seed
        pop_type     = 'campus',
        start_day = '2020-08-31',
        end_day = '2020-11-24'
    )
    pars.interventions = [cvic.symptomQuarantine(symp_prob= 0.5, #This indicates the expected time to find a symptomatic individual is one day
                                                ili_prev=None,
                                                test_sensitivity= 0.95, #Based on a nasopharyngeal swab (Hanson et al. 2020)
                                                test_delay=1, 
                                                start_day=0, 
                                                end_day=None),
                            cv.contact_tracing(trace_probs = {'r':1.,'b':0.75,'f':0.75,'c':0.75}, 
                                                trace_time = 2, 
                                                start_day  = 0, 
                                                end_day    = None, 
                                                presumptive=False),
                            cvic.TestScheduler(sampleGenerator  = residGenerator_T.create,
                                               schedule         = 7,
                                               start_date       = 1,
                                               end_date         = None,
                                               test_sensitivity = 0.95,
                                               test_delay       = 1, 
                                               loss_prob        = 0., 
                                               end_quarantine   = False),
                            cvic.TestScheduler(sampleGenerator  = residGenerator_W.create,
                                               schedule         = 7,
                                               start_date       = 2,
                                               end_date         = None,
                                               test_sensitivity = 0.95,
                                               test_delay       = 1, 
                                               loss_prob        = 0., 
                                               end_quarantine   = False),
                            cvic.TestScheduler(sampleGenerator  = nonResidGenerator.create,
                                               schedule         = 7,
                                               start_date       = 1,
                                               end_date         = None,
                                               test_sensitivity = 0.95,
                                               test_delay       = 1, 
                                               loss_prob        = 0., 
                                               end_quarantine   = False),
                            cvic.TestScheduler(sampleGenerator  = nonResidGenerator.create,
                                               schedule         = 7,
                                               start_date       = 2,
                                               end_date         = None,
                                               test_sensitivity = 0.95,
                                               test_delay       = 1, 
                                               loss_prob        = 0., 
                                               end_quarantine   = False)]

elif scenario == "R0_2.5_Contacts_10_100percent":
    simGroup = "BhartiUpperBound"
    np.random.seed(219222689)
    tuesdayPool   = np.random.choice(7425,3712,replace = False)
    wednesdayPool = np.array([i for i in range(7425) if not i in tuesdayPool])


    dorm_set = "nq_2020_07_24Dedense"
    numberRuns = 1000

    currentNonRes = 7425 - 3636
    nonResImportRate = 13.80490/7
    pars = sc.objdict(
        beta         = 0.030888,
        pop_infected = 0,       # Number of initial infections
        n_imports    = 2.207910/7,
        rand_seed    = 3,        # Random seed
        pop_type     = 'campus',
        start_day = '2020-08-31',
        end_day = '2020-11-24'
    )
    pars.interventions = [cvic.symptomQuarantine(symp_prob= 0.5, #This indicates the expected time to find a symptomatic individual is one day
                                                ili_prev=None,
                                                test_sensitivity= 0.95, #Based on a nasopharyngeal swab (Hanson et al. 2020)
                                                test_delay=1, 
                                                start_day=0, 
                                                end_day=None),
                            cv.contact_tracing(trace_probs = {'r':1.,'b':0.75,'f':0.75,'c':0.75}, 
                                                trace_time = 2, 
                                                start_day  = 0, 
                                                end_day    = None, 
                                                presumptive=False),
                            cvic.TestScheduler(sampleGenerator  = tuesdayPool,
                                               schedule         = 7,
                                               start_date       = 1,
                                               end_date         = None,
                                               test_sensitivity = 0.95,
                                               test_delay       = 1, 
                                               loss_prob        = 0., 
                                               end_quarantine   = False),
                            cvic.TestScheduler(sampleGenerator  = wednesdayPool,
                                               schedule         = 7,
                                               start_date       = 2,
                                               end_date         = None,
                                               test_sensitivity = 0.95,
                                               test_delay       = 1, 
                                               loss_prob        = 0., 
                                               end_quarantine   = False)]

elif scenario == "Holdout_AllInterventions_Targeted":
    simGroup = "BhartiUpperBound"
    residGeneratorAssure = cvic.FloorTargetedPools(0.075,assureCoverage = True,skipQuarantine = True,skipDiagnosed = True,collapse = True,lock = False)
    residGeneratorNotAssure = cvic.FloorTargetedPools(0.075,assureCoverage = False,skipQuarantine = True,skipDiagnosed = True,collapse = True,lock = False)
    

    dorm_set = "nq_2020_07_24Dedense"
    numberRuns = 1000
    currentNonRes = 0
    nonResImportRate = 0/7
    pars = sc.objdict(
        beta         = 0.030888,
        pop_infected = 0,       # Number of initial infections
        n_imports    = 2.207910/7,
        rand_seed    = 3,        # Random seed
        pop_type     = 'campus',
        start_day = '2020-08-31',
        end_day = '2020-11-24'
    )
    pars.interventions = [cvic.symptomQuarantine(symp_prob= 0.5, #This indicates the expected time to find a symptomatic individual is one day
                                                ili_prev=None,
                                                test_sensitivity= 0.95, #Based on a nasopharyngeal swab (Hanson et al. 2020)
                                                test_delay=1, 
                                                start_day=0, 
                                                end_day=None),
                            cv.contact_tracing(trace_probs = {'r':1.,'b':0.75,'f':0.75,'c':0.75}, 
                                                trace_time = 2, 
                                                start_day  = 0, 
                                                end_day    = None, 
                                                presumptive=False),
                            cvic.TestScheduler(sampleGenerator  = residGeneratorAssure.create,
                                               schedule         = 7,
                                               start_date       = 1,
                                               end_date         = None,
                                               test_sensitivity = 0.95,
                                               test_delay       = 1, 
                                               loss_prob        = 0., 
                                               end_quarantine   = False),
                            cvic.TestScheduler(sampleGenerator  = residGeneratorNotAssure.create,
                                               schedule         = 7,
                                               start_date       = 2,
                                               end_date         = None,
                                               test_sensitivity = 0.95,
                                               test_delay       = 1, 
                                               loss_prob        = 0., 
                                               end_quarantine   = False)]

elif scenario == "Holdout_AllInterventions_SRS":
    class RandomResidentSample_SetTests:
        def __init__(self,numberTests,lock = False):
            self.numberTests  = numberTests
            self.lock         = lock
            self.sample       = np.array([])

        def create(self,sim):
            if not (self.lock and len(self.sample) > 0):
                resPop  = sim.dorm_offsets[-1]
                self.sample   = np.random.choice(resPop,self.numberTests,replace = False)
            assert all(self.sample < resPop)
            return self.sample


    simGroup = "BhartiUpperBound"
    residGenerator_T = RandomResidentSample_SetTests(355)
    residGenerator_W = RandomResidentSample_SetTests(225)
    nonResidGenerator = cvic.RandomNonResidentSample(0.125)


    dorm_set = "nq_2020_07_24Dedense"
    numberRuns = 1000
    currentNonRes = 0
    nonResImportRate = 0/7
    pars = sc.objdict(
        beta         = 0.030888,
        pop_infected = 0,       # Number of initial infections
        n_imports    = 2.207910/7,
        rand_seed    = 3,        # Random seed
        pop_type     = 'campus',
        start_day = '2020-08-31',
        end_day = '2020-11-24'
    )
    pars.interventions = [cvic.symptomQuarantine(symp_prob= 0.5, #This indicates the expected time to find a symptomatic individual is one day
                                                ili_prev=None,
                                                test_sensitivity= 0.95, #Based on a nasopharyngeal swab (Hanson et al. 2020)
                                                test_delay=1, 
                                                start_day=0, 
                                                end_day=None),
                            cv.contact_tracing(trace_probs = {'r':1.,'b':0.75,'f':0.75,'c':0.75}, 
                                                trace_time = 2, 
                                                start_day  = 0, 
                                                end_day    = None, 
                                                presumptive=False),
                            cvic.TestScheduler(sampleGenerator  = residGenerator_T.create,
                                               schedule         = 7,
                                               start_date       = 1,
                                               end_date         = None,
                                               test_sensitivity = 0.95,
                                               test_delay       = 1, 
                                               loss_prob        = 0., 
                                               end_quarantine   = False),
                            cvic.TestScheduler(sampleGenerator  = residGenerator_W.create,
                                               schedule         = 7,
                                               start_date       = 2,
                                               end_date         = None,
                                               test_sensitivity = 0.95,
                                               test_delay       = 1, 
                                               loss_prob        = 0., 
                                               end_quarantine   = False),
                            cvic.TestScheduler(sampleGenerator  = nonResidGenerator.create,
                                               schedule         = 7,
                                               start_date       = 1,
                                               end_date         = None,
                                               test_sensitivity = 0.95,
                                               test_delay       = 1, 
                                               loss_prob        = 0., 
                                               end_quarantine   = False),
                            cvic.TestScheduler(sampleGenerator  = nonResidGenerator.create,
                                               schedule         = 7,
                                               start_date       = 2,
                                               end_date         = None,
                                               test_sensitivity = 0.95,
                                               test_delay       = 1, 
                                               loss_prob        = 0., 
                                               end_quarantine   = False)]

elif scenario == "Holdout_DelayedSymptomTesting_Targeted":
    simGroup = "BhartiUpperBound"
    residGeneratorAssure = cvic.FloorTargetedPools(0.075,assureCoverage = True,skipQuarantine = True,skipDiagnosed = True,collapse = True,lock = False)
    residGeneratorNotAssure = cvic.FloorTargetedPools(0.075,assureCoverage = False,skipQuarantine = True,skipDiagnosed = True,collapse = True,lock = False)

    

    dorm_set = "nq_2020_07_24Dedense"
    numberRuns = 1000
    currentNonRes = 0
    nonResImportRate = 0/7
    pars = sc.objdict(
        beta         = 0.030888,
        pop_infected = 0,       # Number of initial infections
        n_imports    = 2.207910/7,
        rand_seed    = 3,        # Random seed
        pop_type     = 'campus',
        start_day = '2020-08-31',
        end_day = '2020-11-24'
    )
    pars.interventions = [cvic.symptomQuarantine(symp_prob= 1./4.82, #This is equivalent to Khalili et al. (2020)'s mean waiting time from symptom onset to first clinic visit
                                                ili_prev=None,
                                                test_sensitivity= 0.95, #Based on a nasopharyngeal swab (Hanson et al. 2020)
                                                test_delay=1, 
                                                start_day=0, 
                                                end_day=None),
                            cv.contact_tracing(trace_probs = {'r':1.,'b':0.75,'f':0.75,'c':0.75}, 
                                                trace_time = 2, 
                                                start_day  = 0, 
                                                end_day    = None, 
                                                presumptive=False),
                            cvic.TestScheduler(sampleGenerator  = residGeneratorAssure.create,
                                               schedule         = 7,
                                               start_date       = 1,
                                               end_date         = None,
                                               test_sensitivity = 0.95,
                                               test_delay       = 1, 
                                               loss_prob        = 0., 
                                               end_quarantine   = False),
                            cvic.TestScheduler(sampleGenerator  = residGeneratorNotAssure.create,
                                               schedule         = 7,
                                               start_date       = 2,
                                               end_date         = None,
                                               test_sensitivity = 0.95,
                                               test_delay       = 1, 
                                               loss_prob        = 0., 
                                               end_quarantine   = False)]

elif scenario == "Holdout_DelayedSymptomTesting_SRS":
    class RandomResidentSample_SetTests:
        def __init__(self,numberTests,lock = False):
            self.numberTests  = numberTests
            self.lock         = lock
            self.sample       = np.array([])

        def create(self,sim):
            if not (self.lock and len(self.sample) > 0):
                resPop  = sim.dorm_offsets[-1]
                self.sample   = np.random.choice(resPop,self.numberTests,replace = False)
            assert all(self.sample < resPop)
            return self.sample


    simGroup = "BhartiUpperBound"
    residGenerator_T = RandomResidentSample_SetTests(355)
    residGenerator_W = RandomResidentSample_SetTests(225)


    dorm_set = "nq_2020_07_24Dedense"
    numberRuns = 1000
    currentNonRes = 0
    nonResImportRate = 0/7
    pars = sc.objdict(
        beta         = 0.030888,
        pop_infected = 0,       # Number of initial infections
        n_imports    = 2.207910/7,
        rand_seed    = 3,        # Random seed
        pop_type     = 'campus',
        start_day = '2020-08-31',
        end_day = '2020-11-24'
    )
    pars.interventions = [cvic.symptomQuarantine(symp_prob= 1./4.82, #This is equivalent to Khalili et al. (2020)'s mean waiting time from symptom onset to first clinic visit
                                                ili_prev=None,
                                                test_sensitivity= 0.95, #Based on a nasopharyngeal swab (Hanson et al. 2020)
                                                test_delay=1, 
                                                start_day=0, 
                                                end_day=None),
                            cv.contact_tracing(trace_probs = {'r':1.,'b':0.75,'f':0.75,'c':0.75}, 
                                                trace_time = 2, 
                                                start_day  = 0, 
                                                end_day    = None, 
                                                presumptive=False),
                            cvic.TestScheduler(sampleGenerator  = residGenerator_T.create,
                                               schedule         = 7,
                                               start_date       = 1,
                                               end_date         = None,
                                               test_sensitivity = 0.95,
                                               test_delay       = 1, 
                                               loss_prob        = 0., 
                                               end_quarantine   = False),
                            cvic.TestScheduler(sampleGenerator  = residGenerator_W.create,
                                               schedule         = 7,
                                               start_date       = 2,
                                               end_date         = None,
                                               test_sensitivity = 0.95,
                                               test_delay       = 1, 
                                               loss_prob        = 0., 
                                               end_quarantine   = False)]

elif scenario == "Holdout_noSymptomTesting_Targeted":
    simGroup = "BhartiUpperBound"
    residGeneratorAssure = cvic.FloorTargetedPools(0.075,assureCoverage = True,skipQuarantine = True,skipDiagnosed = True,collapse = True,lock = False)
    residGeneratorNotAssure = cvic.FloorTargetedPools(0.075,assureCoverage = False,skipQuarantine = True,skipDiagnosed = True,collapse = True,lock = False)
    

    dorm_set = "nq_2020_07_24Dedense"
    numberRuns = 1000
    currentNonRes = 0
    nonResImportRate = 0/7
    pars = sc.objdict(
        beta         = 0.030888,
        pop_infected = 0,       # Number of initial infections
        n_imports    = 2.207910/7,
        rand_seed    = 3,        # Random seed
        pop_type     = 'campus',
        start_day = '2020-08-31',
        end_day = '2020-11-24'
    )
    pars.interventions = [cv.contact_tracing(trace_probs = {'r':1.,'b':0.75,'f':0.75,'c':0.75}, 
                                                trace_time = 2, 
                                                start_day  = 0, 
                                                end_day    = None, 
                                                presumptive=False),
                            cvic.TestScheduler(sampleGenerator  = residGeneratorAssure.create,
                                               schedule         = 7,
                                               start_date       = 1,
                                               end_date         = None,
                                               test_sensitivity = 0.95,
                                               test_delay       = 1, 
                                               loss_prob        = 0., 
                                               end_quarantine   = False),
                            cvic.TestScheduler(sampleGenerator  = residGeneratorNotAssure.create,
                                               schedule         = 7,
                                               start_date       = 2,
                                               end_date         = None,
                                               test_sensitivity = 0.95,
                                               test_delay       = 1, 
                                               loss_prob        = 0., 
                                               end_quarantine   = False)]

elif scenario == "Holdout_noSymptomTesting_SRS":
    class RandomResidentSample_SetTests:
        def __init__(self,numberTests,lock = False):
            self.numberTests  = numberTests
            self.lock         = lock
            self.sample       = np.array([])

        def create(self,sim):
            if not (self.lock and len(self.sample) > 0):
                resPop  = sim.dorm_offsets[-1]
                self.sample   = np.random.choice(resPop,self.numberTests,replace = False)
            assert all(self.sample < resPop)
            return self.sample


    simGroup = "BhartiUpperBound"
    residGenerator_T = RandomResidentSample_SetTests(355)
    residGenerator_W = RandomResidentSample_SetTests(225)


    dorm_set = "nq_2020_07_24Dedense"
    numberRuns = 1000
    currentNonRes = 0
    nonResImportRate = 0/7
    pars = sc.objdict(
        beta         = 0.030888,
        pop_infected = 0,       # Number of initial infections
        n_imports    = 2.207910/7,
        rand_seed    = 3,        # Random seed
        pop_type     = 'campus',
        start_day = '2020-08-31',
        end_day = '2020-11-24'
    )
    pars.interventions = [cv.contact_tracing(trace_probs = {'r':1.,'b':0.75,'f':0.75,'c':0.75}, 
                                                trace_time = 2, 
                                                start_day  = 0, 
                                                end_day    = None, 
                                                presumptive=False),
                            cvic.TestScheduler(sampleGenerator  = residGenerator_T.create,
                                               schedule         = 7,
                                               start_date       = 1,
                                               end_date         = None,
                                               test_sensitivity = 0.95,
                                               test_delay       = 1, 
                                               loss_prob        = 0., 
                                               end_quarantine   = False),
                            cvic.TestScheduler(sampleGenerator  = residGenerator_W.create,
                                               schedule         = 7,
                                               start_date       = 2,
                                               end_date         = None,
                                               test_sensitivity = 0.95,
                                               test_delay       = 1, 
                                               loss_prob        = 0., 
                                               end_quarantine   = False)]

elif scenario == "Holdout_noContactTracing_Targeted":
    simGroup = "BhartiUpperBound"
    residGeneratorAssure = cvic.FloorTargetedPools(0.075,assureCoverage = True,skipQuarantine = True,skipDiagnosed = True,collapse = True,lock = False)
    residGeneratorNotAssure = cvic.FloorTargetedPools(0.075,assureCoverage = False,skipQuarantine = True,skipDiagnosed = True,collapse = True,lock = False)
     

    dorm_set = "nq_2020_07_24Dedense"
    numberRuns = 1000
    currentNonRes = 0
    nonResImportRate = 0/7
    pars = sc.objdict(
        beta         = 0.030888,
        pop_infected = 0,       # Number of initial infections
        n_imports    = 2.207910/7,
        rand_seed    = 3,        # Random seed
        pop_type     = 'campus',
        start_day = '2020-08-31',
        end_day = '2020-11-24'
    )
    pars.interventions = [cvic.symptomQuarantine(symp_prob= 0.5, #This indicates the expected time to find a symptomatic individual is one day
                                                ili_prev=None,
                                                test_sensitivity= 0.95, #Based on a nasopharyngeal swab (Hanson et al. 2020)
                                                test_delay=1, 
                                                start_day=0, 
                                                end_day=None),
                            cvic.TestScheduler(sampleGenerator  = residGeneratorAssure.create,
                                               schedule         = 7,
                                               start_date       = 1,
                                               end_date         = None,
                                               test_sensitivity = 0.95,
                                               test_delay       = 1, 
                                               loss_prob        = 0., 
                                               end_quarantine   = False),
                            cvic.TestScheduler(sampleGenerator  = residGeneratorNotAssure.create,
                                               schedule         = 7,
                                               start_date       = 2,
                                               end_date         = None,
                                               test_sensitivity = 0.95,
                                               test_delay       = 1, 
                                               loss_prob        = 0., 
                                               end_quarantine   = False)]


elif scenario == "Holdout_noContactTracing_SRS":
    class RandomResidentSample_SetTests:
        def __init__(self,numberTests,lock = False):
            self.numberTests  = numberTests
            self.lock         = lock
            self.sample       = np.array([])

        def create(self,sim):
            if not (self.lock and len(self.sample) > 0):
                resPop  = sim.dorm_offsets[-1]
                self.sample   = np.random.choice(resPop,self.numberTests,replace = False)
            assert all(self.sample < resPop)
            return self.sample


    simGroup = "BhartiUpperBound"
    residGenerator_T = RandomResidentSample_SetTests(355)
    residGenerator_W = RandomResidentSample_SetTests(225)


    dorm_set = "nq_2020_07_24Dedense"
    numberRuns = 1000
    currentNonRes = 0
    nonResImportRate = 0/7
    pars = sc.objdict(
        beta         = 0.030888,
        pop_infected = 0,       # Number of initial infections
        n_imports    = 2.207910/7,
        rand_seed    = 3,        # Random seed
        pop_type     = 'campus',
        start_day = '2020-08-31',
        end_day = '2020-11-24'
    )
    pars.interventions = [cvic.symptomQuarantine(symp_prob= 0.5, #This indicates the expected time to find a symptomatic individual is one day
                                                ili_prev=None,
                                                test_sensitivity= 0.95, #Based on a nasopharyngeal swab (Hanson et al. 2020)
                                                test_delay=1, 
                                                start_day=0, 
                                                end_day=None),
                            cvic.TestScheduler(sampleGenerator  = residGenerator_T.create,
                                               schedule         = 7,
                                               start_date       = 1,
                                               end_date         = None,
                                               test_sensitivity = 0.95,
                                               test_delay       = 1, 
                                               loss_prob        = 0., 
                                               end_quarantine   = False),
                            cvic.TestScheduler(sampleGenerator  = residGenerator_W.create,
                                               schedule         = 7,
                                               start_date       = 2,
                                               end_date         = None,
                                               test_sensitivity = 0.95,
                                               test_delay       = 1, 
                                               loss_prob        = 0., 
                                               end_quarantine   = False)]

else:
    raise ValueError("Unknown scenario encountered. Please try again.")


if simGroup == "BhartiUpperBound":
    currentSim = cvsc.SimCampus(pars=pars,dorms = qud.get_QU_Dorms(dorm_set),contacts = {'r': -1,'b': 1.5,'f': 1,'c': 2},nonResident = currentNonRes,n_importsNonRes = nonResImportRate)
elif simGroup == "new8ContactGroup":
    currentSim = cvsc.SimCampus(pars=pars,dorms = qud.get_QU_Dorms(dorm_set),contacts = {'r': -1,'b': 1.5,'f': 1,'c': 1.25},nonResident = currentNonRes,n_importsNonRes = nonResImportRate)
else:
    raise ValueError("Unknown simGroup encountered. Please try again.")


if __name__ == '__main__':
    freeze_support()
    sim = currentSim
    multiSim = cv.MultiSim(sims = currentSim,n_runs =numberRuns,quantiles = {'low':0.05,'high':0.95})
    multiSim.run(verbose=0)
    multiSim.reduce()
    endTime = datetime.now()

    print('\n' + str(multiSim.summary))
    print('\nQuantiles:')
    print('Cumulative Case: [' + str(multiSim.results['cum_infections'].low[-1]) + ", " + str(multiSim.results['cum_infections'].high[-1]) + "]")
    print('Cumulative Severe: [' + str(multiSim.results['cum_severe'].low[-1]) + ", " + str(multiSim.results['cum_severe'].high[-1]) + "]")
    print('Cumulative Critical: [' + str(multiSim.results['cum_critical'].low[-1]) + ", " + str(multiSim.results['cum_critical'].high[-1]) + "]")
    print('Cumulative Deaths: [' + str(multiSim.results['cum_deaths'].low[-1]) + ", " + str(multiSim.results['cum_deaths'].high[-1]) + "]")
    print('\nExecution Time: ' + str(endTime - startTime))
    if writeInfectionToFile:
        DataFrame({'cum_infects_low': multiSim.results['cum_infections'].low,
                   'cum_infects_med': multiSim.results['cum_infections'].values,
                   'cum_infects_high': multiSim.results['cum_infections'].high}).to_csv(seriesFilePath)
        DataFrame({'cum_diagnoses_low': multiSim.results['cum_diagnoses'].low,
                   'cum_diagnoses_med': multiSim.results['cum_diagnoses'].values,
                   'cum_diagnoses_high': multiSim.results['cum_diagnoses'].high}).to_csv(diagnosedFilePath)        
        DataFrame({'cum_tests_low': multiSim.results['cum_tests'].low,
                   'cum_tests_med': multiSim.results['cum_tests'].values,
                   'cum_tests_high': multiSim.results['cum_tests'].high}).to_csv(testsFilePath)

    experimentPlot = {'Quarantine':['n_quarantineDorm','n_quarantined'],'tests':['new_tests']}
    multiSim.plot(sep_figs = True,dateformat = "%m/%d")
    multiSim.plot(sep_figs = True,to_plot = experimentPlot)
```
