## Supplementary material for "Monitoring and responding to emerging infectious diseases in a university setting: A case study using COVID-19": S2 Supporting Information: QUDormsLegacy_20200904_208.py

```
import covasim.base as cvb
import covasim.baseCampus as cvbc

import math
import numpy as np
import pandas as pd
import random as rd


def get_QU_Dorms(desiredSet = "all"):
    if desiredSet == "all":
        currentDorms = cvb.FlexDict({'DanaEnglish':cvbc.Dorm(2,[10,15,15],[2,2,2]),
                            'MountainView':cvbc.Dorm(2,[4,4,4,4,4],[8,14,14,5,5]),
                            'Commons':cvbc.Dorm(4,[13,13,13],[3,3,3]),
                            'Ledges': cvbc.Dorm(4,[5,15,16,16],[1,2,2,2]),
                            'Irmagarde-Tator': cvbc.Dorm(2,[10,15,15],[2,2,2]),
                            'Larson': cvbc.Dorm(2,[4,4],[8,10]),
                            'Perlroth': cvbc.Dorm(2,[4,4],[8,10]),
                            'Troup': cvbc.Dorm(2,[4,4],[8,10]),
                            'Sahlin': cvbc.Dorm(2,[3,3,3],[2,4,4]),
                            'Bakke': cvbc.Dorm(2,[3,3],[4,4]),
                            'Founders': cvbc.Dorm(2,[3,3,3],[3,4,4]),
                            'HillG': cvbc.Dorm(2,[3,3,3],[8,8,8]),
                            'HillF': cvbc.Dorm(2,[3,3,3],[6,6,6]),
                            'Crescent': cvbc.Dorm(2,[3,3,3,3,3,3,3,3,3],[5,14,18,22,21,21,11,5,3]), 
                            'Westview': cvbc.FlexDorm([[[1,1,1,2]],[[1,1,1,2]],[[1,1,1,2]],[[1,1,1,2]],[[1,1,1,2]],[[1,1,1,2]],[[1,1,1,2]]],[[4],[7],[8],[8],[8],[8],[4]]), 
                            'Eastview': cvbc.FlexDorm([[[1,1,1,2]],[[1,1,1,2],[1,1,1,1,2]],[[1,1,1,2],[1,1,1,1,2]],[[1,1,1,2],[1,1,1,1,2]],[[1,1,1,2],[1,1,1,1,2]],[[1,1,1,2],[1,1,1,1,2]],[[1,1,1,2],[1,1,1,1,2]]],[[5],[4,1],[5,1],[5,1],[5,1],[1,3],[1,1]]),  #This one is the most problematic b/c the number of people per room/suite is highly heterogeneous
                            'TownhouseExtraFloor:': cvbc.Dorm(2,[3,3,3],[1,1,1]) 
                             })
        currentDorms.update(cvbc.autoCreateDorms(28,2,[3,3],[1,1],"Village"))

        currentDorms.update(cvbc.autoCreateDorms(9,2,[3,3],[1,1],"Townhouses")) 

        currentDorms.update(cvbc.autoCreateDorms(3,2,[1],[1],"2BDHouse")) 
        currentDorms.update(cvbc.autoCreateDorms(29,3,[1],[1],"3BDHouse")) 
        currentDorms.update(cvbc.autoCreateDorms(21,4,[1],[1],"4BDHouse")) 


        

    elif desiredSet == "nonQuarantined":
        currentDorms = cvb.FlexDict({'DanaEnglish':cvbc.Dorm(2,[10,15,15],[2,2,2]),
                            'MountainView':cvbc.Dorm(2,[4,4,4,4,4],[8,14,14,5,5]),
                            'Commons':cvbc.Dorm(4,[13,13,13],[3,3,3]),
                            'Ledges': cvbc.Dorm(4,[5,15,16,16],[1,2,2,2]),
                            'Irmagarde-Tator': cvbc.Dorm(2,[10,15,15],[2,2,2]),
                            'Larson': cvbc.Dorm(2,[4,4],[8,10]),
                            'Perlroth': cvbc.Dorm(2,[4,4],[8,10]),
                            'Troup': cvbc.Dorm(2,[4,4],[8,10]),
                            'Sahlin': cvbc.Dorm(2,[3,3,3],[2,4,4]),
                            'HillG': cvbc.Dorm(2,[3,3,3],[8,8,8]),
                            'HillF': cvbc.Dorm(2,[3,3,3],[6,6,6]),
                            'Crescent': cvbc.Dorm(2,[3,3,3,3,3,3,3,3,3],[5,14,18,22,21,21,11,5,3]), 
                            'Westview': cvbc.FlexDorm([[[1,1,1,2]],[[1,1,1,2]],[[1,1,1,2]],[[1,1,1,2]],[[1,1,1,2]],[[1,1,1,2]],[[1,1,1,2]]],[[4],[7],[8],[8],[8],[8],[4]]), 
                            'Eastview': cvbc.FlexDorm([[[1,1,1,2]],[[1,1,1,2],[1,1,1,1,2]],[[1,1,1,2],[1,1,1,1,2]],[[1,1,1,2],[1,1,1,1,2]],[[1,1,1,2],[1,1,1,1,2]],[[1,1,1,2],[1,1,1,1,2]],[[1,1,1,2],[1,1,1,1,2]]],[[5],[4,1],[5,1],[5,1],[5,1],[1,3],[1,1]]),  
                            'TownhouseExtraFloor:': cvbc.Dorm(2,[3,3,3],[1,1,1]) 
                            })
        currentDorms.update(cvbc.autoCreateDorms(28,2,[3,3],[1,1],"Village")) 

        currentDorms.update(cvbc.autoCreateDorms(9,2,[3,3],[1,1],"Townhouses")) 

        currentDorms.update(cvbc.autoCreateDorms(3,2,[1],[1],"2BDHouse")) 
        currentDorms.update(cvbc.autoCreateDorms(29,3,[1],[1],"3BDHouse"))
        currentDorms.update(cvbc.autoCreateDorms(21,4,[1],[1],"4BDHouse")) 

    elif desiredSet == "nq_2020_07_24":
        currentDorms = cvb.FlexDict({'DanaEnglish':cvbc.Dorm(2,[10,15,15],[2,2,2]),
                            'MountainView':cvbc.Dorm(2,[4,4,4,4,4],[8,14,14,5,5]),
                            'Commons':cvbc.Dorm(4,[13,13,13],[3,3,3]),
                            'Ledges': cvbc.Dorm(4,[5,15,16,16],[1,2,2,2]),
                            'Irmagarde-Tator': cvbc.Dorm(2,[10,15,15],[2,2,2]),
                            'Larson': cvbc.Dorm(2,[4,4],[8,10]),
                            'Perlroth': cvbc.Dorm(2,[4,4],[8,10]),
                            'Troup': cvbc.Dorm(2,[4,4],[8,10]),
                            'Sahlin': cvbc.Dorm(2,[3,3,3],[2,4,4]),
                            'HillG': cvbc.Dorm(2,[3,3,3],[8,8,8]),
                            'HillF': cvbc.Dorm(2,[3,3,3],[6,6,6]),
                            'Crescent': cvbc.Dorm(2,[3,3,3,3,3,3,3,3,3],[5,14,18,22,21,21,11,5,3]), 
                            'Westview': cvbc.FlexDorm([[[1,1,1,2]],[[1,1,1,2]],[[1,1,1,2]],[[1,1,1,2]],[[1,1,1,2]],[[1,1,1,2]],[[1,1,1,2]]],[[4],[7],[8],[8],[8],[8],[4]]),
                            'Eastview': cvbc.FlexDorm([[[1,1,1,2]],[[1,1,1,2],[1,1,1,1,2]],[[1,1,1,2],[1,1,1,1,2]],[[1,1,1,2],[1,1,1,1,2]],[[1,1,1,2],[1,1,1,1,2]],[[1,1,1,2],[1,1,1,1,2]],[[1,1,1,2],[1,1,1,1,2]]],[[5],[4,1],[5,1],[5,1],[5,1],[1,3],[1,1]]),
                            'TownhouseExtraFloor:': cvbc.Dorm(2,[3,3,3],[1,1,1]) 
                            })
        currentDorms.update(cvbc.autoCreateDorms(28,2,[3,3],[1,1],"Village")) 

        currentDorms.update(cvbc.autoCreateDorms(9,2,[3,3],[1,1],"Townhouses"))

        currentDorms.update(cvbc.autoCreateDorms(3,2,[1],[1],"2BDHouse")) 

        currentDorms.update(cvbc.autoCreateDorms(29,3,[1],[1],"3BDHouse"))
        currentDorms.update(cvbc.autoCreateDorms(21,4,[1],[1],"4BDHouse")) 

    elif desiredSet == "nq_2020_07_24Dedense_typeSpecify":
        currentDorms = cvb.FlexDict({'DanaEnglish':cvbc.Dorm(2,[10,15,15],[2,2,2],"communal"),
                            'MountainView':cvbc.Dorm(2,[4,4,4,4,4],[8,14,14,5,5],"suite"),
                            'Commons':cvbc.Dorm(2,[13,13,13],[3,3,3],"communal"),
                            'Ledges': cvbc.Dorm(2,[5,15,16,16],[1,2,2,2],"communal"),
                            'Irmagarde-Tator': cvbc.Dorm(2,[10,15,15],[2,2,2],"communal"),
                            'Larson': cvbc.Dorm(2,[4,4],[8,10],"suite"),
                            'Perlroth': cvbc.Dorm(2,[4,4],[8,10],"suite"),
                            'Troup': cvbc.Dorm(2,[4,4],[8,10],"suite"),
                            'Sahlin': cvbc.Dorm(2,[3,3,3],[2,4,4],"suite"),
                            'HillG': cvbc.Dorm(2,[3,3,3],[8,8,8],"suite"),
                            'HillF': cvbc.Dorm(2,[3,3,3],[6,6,6],"suite"),
                            'Crescent': cvbc.Dorm(2,[3,3,3,3,3,3,3,3,3],[5,14,18,22,21,21,11,5,3],"suite"), 
                            'Westview': cvbc.FlexDorm([[[1,1,1,2]],[[1,1,1,2]],[[1,1,1,2]],[[1,1,1,2]],[[1,1,1,2]],[[1,1,1,2]],[[1,1,1,2]]],[[4],[7],[8],[8],[8],[8],[4]],"suite"), 

                            'Eastview': cvbc.FlexDorm([[[1,1,1,2]],[[1,1,1,2],[1,1,1,1,2]],[[1,1,1,2],[1,1,1,1,2]],[[1,1,1,2],[1,1,1,1,2]],[[1,1,1,2],[1,1,1,1,2]],[[1,1,1,2],[1,1,1,1,2]],[[1,1,1,2],[1,1,1,1,2]]],[[5],[4,1],[5,1],[5,1],[5,1],[1,3],[1,1]],"suite"), 
                            'TownhouseExtraFloor:': cvbc.Dorm(2,[3,3,3],[1,1,1],"suite")
                            })
        currentDorms.update(cvbc.autoCreateDorms(28,2,[3,3],[1,1],"Village","suite"))

        currentDorms.update(cvbc.autoCreateDorms(9,2,[3,3],[1,1],"Townhouses","suite")) 

        currentDorms.update(cvbc.autoCreateDorms(3,2,[1],[1],"2BDHouse","suite")) 
        currentDorms.update(cvbc.autoCreateDorms(29,3,[1],[1],"3BDHouse","suite"))
        currentDorms.update(cvbc.autoCreateDorms(21,4,[1],[1],"4BDHouse","suite")) 

    elif desiredSet == "nq_2020_07_24Dedense":
        currentDorms = cvb.FlexDict({'DanaEnglish':cvbc.Dorm(2,[10,15,15],[2,2,2]),
                            'MountainView':cvbc.Dorm(2,[4,4,4,4,4],[8,14,14,5,5]),
                            'Commons':cvbc.Dorm(2,[13,13,13],[3,3,3]),
                            'Ledges': cvbc.Dorm(2,[5,15,16,16],[1,2,2,2]),
                            'Irmagarde-Tator': cvbc.Dorm(2,[10,15,15],[2,2,2]),
                            'Larson': cvbc.Dorm(2,[4,4],[8,10]),
                            'Perlroth': cvbc.Dorm(2,[4,4],[8,10]),
                            'Troup': cvbc.Dorm(2,[4,4],[8,10]),
                            'Sahlin': cvbc.Dorm(2,[3,3,3],[2,4,4]),
                            'HillG': cvbc.Dorm(2,[3,3,3],[8,8,8]),
                            'HillF': cvbc.Dorm(2,[3,3,3],[6,6,6]),
                            'Crescent': cvbc.Dorm(2,[3,3,3,3,3,3,3,3,3],[5,14,18,22,21,21,11,5,3]), 
                            'Westview': cvbc.FlexDorm([[[1,1,1,2]],[[1,1,1,2]],[[1,1,1,2]],[[1,1,1,2]],[[1,1,1,2]],[[1,1,1,2]],[[1,1,1,2]]],[[4],[7],[8],[8],[8],[8],[4]]), 
                            'Eastview': cvbc.FlexDorm([[[1,1,1,2]],[[1,1,1,2],[1,1,1,1,2]],[[1,1,1,2],[1,1,1,1,2]],[[1,1,1,2],[1,1,1,1,2]],[[1,1,1,2],[1,1,1,1,2]],[[1,1,1,2],[1,1,1,1,2]],[[1,1,1,2],[1,1,1,1,2]]],[[5],[4,1],[5,1],[5,1],[5,1],[1,3],[1,1]]),  
                            'TownhouseExtraFloor:': cvbc.Dorm(2,[3,3,3],[1,1,1]) 
                            })
        currentDorms.update(cvbc.autoCreateDorms(28,2,[3,3],[1,1],"Village"))

        currentDorms.update(cvbc.autoCreateDorms(9,2,[3,3],[1,1],"Townhouses"))

        currentDorms.update(cvbc.autoCreateDorms(3,2,[1],[1],"2BDHouse")) 

        currentDorms.update(cvbc.autoCreateDorms(29,3,[1],[1],"3BDHouse"))
        currentDorms.update(cvbc.autoCreateDorms(21,4,[1],[1],"4BDHouse")) 

    elif desiredSet == "nq_2020_07_08":
        currentDorms = cvb.FlexDict({'DanaEnglish':cvbc.Dorm(2,[10,15,15],[2,2,2]),
                            'MountainView':cvbc.Dorm(2,[4,4,4,4,4],[8,14,14,5,5]),
                            'Commons':cvbc.Dorm(4,[13,13,13],[3,3,3]),
                            'Ledges': cvbc.Dorm(4,[5,15,16,16],[1,2,2,2]),
                            'Irmagarde-Tator': cvbc.Dorm(2,[10,15,15],[2,2,2]),
                            'Larson': cvbc.Dorm(2,[4,4],[8,10]),
                            'Perlroth': cvbc.Dorm(2,[4,4],[8,10]),
                            'Troup': cvbc.Dorm(2,[4,4],[8,10]),
                            'Sahlin': cvbc.Dorm(2,[3,3,3],[2,4,4]),
                            'Bakke': cvbc.Dorm(2,[3,3],[4,4]),
                            'Founders': cvbc.Dorm(2,[3,3,3],[3,4,4]),
                            'HillG': cvbc.Dorm(2,[3,3,3],[8,8,8]),
                            'HillF': cvbc.Dorm(2,[3,3,3],[6,6,6])})
        currentDorms.update(cvbc.autoCreateDorms(28,2,[3,3],[1,1],"Village"))
        currentDorms.update(cvbc.autoCreateDorms(3,2,[1],[1],"2BDHouse")) 
        currentDorms.update(cvbc.autoCreateDorms(29,3,[1],[1],"3BDHouse")) 
        currentDorms.update(cvbc.autoCreateDorms(21,4,[1],[1],"4BDHouse")) 
    elif desiredSet == "nq_2020_06_24":
        currentDorms = cvb.FlexDict({'DanaEnglish':cvbc.Dorm(2,[10,15,15],[2,2,2]),
                            'MountainView':cvbc.Dorm(2,[4,4,4,4,4],[8,14,14,5,5]),
                            'Commons':cvbc.Dorm(4,[13,13,13],[3,3,3]),
                            'Ledges': cvbc.Dorm(4,[5,15,16,16],[1,2,2,2]),
                            'Irmagarde-Tator': cvbc.Dorm(2,[10,15,15],[2,2,2]),
                            'Larson': cvbc.Dorm(2,[4,4],[8,10]),
                            'Perlroth': cvbc.Dorm(2,[4,4],[8,10]),
                            'Troup': cvbc.Dorm(2,[4,4],[8,10]),
                            'Sahlin': cvbc.Dorm(2,[3,3,3],[2,4,4]),
                            'HillG': cvbc.Dorm(2,[3,3,3],[8,8,8]),
                            'HillF': cvbc.Dorm(2,[3,3,3],[6,6,6])})
        currentDorms.update(cvbc.autoCreateDorms(28,2,[3,3],[1,1]))
    elif desiredSet == "nq_2020_06_10":
        currentDorms = cvb.FlexDict({'DanaEnglish':cvbc.Dorm(2,[10,15,15],[2,2,2]),
                            'MountainView':cvbc.Dorm(2,[4,4,4,4,4],[8,14,14,5,5]),
                            'Commons':cvbc.Dorm(4,[13,13,13],[3,3,3]),
                            'Ledges': cvbc.Dorm(4,[5,15,16,16],[1,2,2,2]),
                            'Irmagarde-Tator': cvbc.Dorm(2,[10,15,15],[2,2,2]),
                            'Larson': cvbc.Dorm(2,[4,4],[8,10]),
                            'Perlroth': cvbc.Dorm(2,[4,4],[8,10]),
                            'Troup': cvbc.Dorm(2,[4,4],[8,10]),
                            'Sahlin': cvbc.Dorm(2,[3,3,3],[2,4,4]),
                            'HillG': cvbc.Dorm(2,[3,3,3],[8,8,8]),
                            'HillF': cvbc.Dorm(2,[3,3,3],[6,6,6]),
                            'Village': cvbc.Dorm(2,[3,3,3],[6,6,6])})

    elif desiredSet == "2020_06_03":
        currentDorms = cvb.FlexDict({'DanaEnglish':cvbc.Dorm(2,[10,15,15],[2,2,2]),
                            'MountainView':cvbc.Dorm(2,[4,4,4,4,4],[8,14,14,5,5]),
                            'Commons':cvbc.Dorm(4,[13,13,13],[3,3,3])})
    elif desiredSet == "2020_06_08":
        currentDorms = cvb.FlexDict({'DanaEnglish':cvbc.Dorm(2,[10,15,15],[2,2,2]),
                            'MountainView':cvbc.Dorm(2,[4,4,4,4,4],[8,14,14,5,5]),
                            'Commons':cvbc.Dorm(4,[13,13,13],[3,3,3]),
                            'Ledges': cvbc.Dorm(4,[5,15,16,16],[1,2,2,2])})
    else:
        raise RuntimeWarning("Unknown value encountered. Defaulting to 'all'")
        currentDorms = get_QU_Dorms("all")

    return currentDorms


def get_QU_Control_Dorms(desiredSet = "all"):
    if desiredSet == "all":
        currentDorms = cvb.FlexDict({'DanaEnglish':cvbc.Dorm(1,[20,30,30],[2,2,2]),
                            'MountainView':cvbc.Dorm(1,[8,8,8,8,8],[8,14,14,5,5]),
                            'Commons':cvbc.Dorm(1,[52,52,52],[3,3,3])})
    elif desiredSet == "2020_06_03":
        currentDorms = cvb.FlexDict({'DanaEnglish':cvbc.Dorm(1,[20,30,30],[2,2,2]),
                            'MountainView':cvbc.Dorm(1,[8,8,8,8,8],[8,14,14,5,5]),
                            'Commons':cvbc.Dorm(1,[52,52,52],[3,3,3])})
    else:
        raise RuntimeWarning("Unknown value encountered. Defaulting to 'all'")
        currentDorms = get_QU_Dorms("all")

    return currentDorms


def get_QU_Dorm_Population(desiredSet):
    if desiredSet == "2020_06_03":
        currentPop = 996
    elif desiredSet == "nq_2020_06_10":
        currentPop = 2404
    else:
        raise ValueError("QUDorms::get_QU_Dorm_Population encountered an unknown value.")


    return currentPop

def get_Number_Floor_Targeted_Samples(desiredSet,coverage,assureCoverage = True):
    dorms = get_QU_Dorms(desiredSet)
    total = 0
    for key, val in dorms.items():
        samples = 0
        for floor in set(val['f']):
            temp = round(sum(val['f'] == floor) * coverage) 
            if assureCoverage and temp == 0:
                temp = 1
            samples += temp
            total   += temp
        print(key + ": " + str(samples) + " tests")
    print("\nTotal: " + str(total) + " tests")
    return

def parseDormStr(dormStr):
    counter = 0
    while not (dormStr[counter]).isnumeric():
        counter += 1
    dormCode = dormStr[:counter]

    strBreak = counter
    while counter < len(dormStr) and (dormStr[counter]).isnumeric():
        counter += 1
    bathroomCode = dormStr[strBreak:counter]

    if counter < len(dormStr):
        roomCode = dormStr[counter:]
    else:
        roomCode = None

    return dormCode, bathroomCode, roomCode 


def extractDormInfo(dormTable):
    processedDorms = pd.DataFrame({'dorm':np.array([],dtype = 'U'),
                                   'floor':np.array([],dtype = int),
                                   'bathroom':np.array([],dtype = int),
                                   'room':np.array([],dtype = 'U'),
                                   'occupancy':np.array([],dtype = int)})

    for i in dormTable.index:
        dormCode, bathroomCode, roomCode = parseDormStr(dormTable['room'][i])
        if roomCode:
            floorToken    = int(bathroomCode[0])
            bathroomToken = int(bathroomCode)
            roomToken     = bathroomCode + roomCode
            if processedDorms.loc[(processedDorms['dorm'] == dormCode) & (processedDorms['floor'] == floorToken) & (processedDorms['bathroom'] == bathroomToken) & (processedDorms['room'] == roomToken)].shape[0] > 0:
                processedDorms.loc[(processedDorms['dorm'] == dormCode) & (processedDorms['floor'] == floorToken) & (processedDorms['bathroom'] == bathroomToken) & (processedDorms['room'] == roomToken),'occupancy'] += 1
            else:
                processedDorms = processedDorms.append(pd.DataFrame({'dorm':np.array([dormCode]),
                                                                     'floor':np.array([floorToken]),
                                                                     'bathroom':np.array([bathroomToken]),
                                                                     'room':np.array([roomToken]),
                                                                     'occupancy':np.array([1])}),ignore_index = True)
        else:
            floorToken    = int(bathroomCode[0])
            bathroomToken = None
            roomToken     = bathroomCode
            if processedDorms.loc[(processedDorms['dorm'] == dormCode) & (processedDorms['floor'] == floorToken) & (processedDorms['room'] == roomToken)].shape[0] > 0:
                processedDorms.loc[(processedDorms['dorm'] == dormCode) & (processedDorms['floor'] == floorToken) & (processedDorms['room'] == roomToken),'occupancy'] += 1
            else:
                processedDorms = processedDorms.append(pd.DataFrame({'dorm':np.array([dormCode]),
                                                                     'floor':np.array([floorToken]),
                                                                     'bathroom':np.array([bathroomToken]),
                                                                     'room':np.array([roomToken]),
                                                                     'occupancy':np.array([1])}),ignore_index = True)

    return processedDorms


'''
    This function will add a column to a pandas.DataFrame containing housing data in preparation for sampling.
'''

def setupSampling(dormTable):
    dormTable['samplingUnit'] = [None] * dormTable.shape[0]
    dormTable['floorID'] = [None] * dormTable.shape[0]

    for i in dormTable.index:
        print(i)
        if isinstance(dormTable['Building'][i],str) and (isinstance(dormTable['Floor'][i],int) or isinstance(dormTable['Floor'][i],np.int64) or isinstance(dormTable['Floor'][i],float)):
            dormTable.loc[i,dormTable.columns == 'floorID'] = dormTable['Building'][i] + str(int(dormTable['Floor'][i]))
        elif isinstance(dormTable['Building'][i],float) and math.isnan(dormTable['Building'][i]) and isinstance(dormTable['Floor'][i],float) and math.isnan(dormTable['Floor'][i]):
            dormTable.loc[i,dormTable.columns == 'floorID'] = "nonQUResident"
        else:
            raise ValueError("No floorID coding instructions were found for this entry. Check entry: " + str(i))

        if isinstance(dormTable['Suite'][i],str):
            dormTable.loc[i,dormTable.columns == 'samplingUnit'] = dormTable['Suite'][i]
        else:
            if dormTable['Building'][i] in ["Crescent","Eastview","Troup Hall"]:
                dormCode, bathroomCode, roomCode = parseDormStr(dormTable['Room'][i])
                dormTable.loc[i,dormTable.columns == 'samplingUnit'] = bathroomCode
            elif dormTable['Building'][i] in ["Dana English Hall","Commons","Irmagarde Tator Hall","Ledges","Mountainview","Whitney Village"]:
                dormTable.loc[i,dormTable.columns == 'samplingUnit'] = dormTable['Room'][i]
            elif dormTable['Building'][i] == "The Village":
                dormTable.loc[i,dormTable.columns == 'samplingUnit'] = str(dormTable['Room'][i])[:3]
            elif isinstance(dormTable['Building'][i],str):
                dormTable.loc[i,dormTable.columns == 'samplingUnit'] = dormTable['Building'][i]
            else:
                dormTable.loc[i,dormTable.columns == 'samplingUnit'] = dormTable['Local Addr 1'][i]

    return dormTable


def generateSample(preppedDormFile,samplingPercent,seed = 0,outputFile = None,floorKey = "fl"):
    allFloors   = set(preppedDormFile[floorKey])
    sampleFrame = pd.DataFrame()

    for floor in allFloors:
        print("Floor Code: " + floor)
        floorFrame     = preppedDormFile.loc[preppedDormFile[floorKey] == floor]
        neededSampleN = round(floorFrame.shape[0] * samplingPercent)
        if neededSampleN == 0:
            neededSampleN = 1
        print("neededSampleN: " + str(neededSampleN))
        possibleUnits  = list(set(floorFrame['samplingUnit']))
        sampledUnits = possibleUnits * int(neededSampleN/len(possibleUnits))
        sampledUnits += rd.sample(possibleUnits,neededSampleN % len(possibleUnits))
        print("sampledUnits: " + str(sampledUnits) + '\n')
        processedSample = validateProposedSample(np.array(sampledUnits),floorFrame,"samplingUnit")
        while (processedSample["Slack"] < 0).any():
            shiftRequired = processedSample[processedSample["Slack"] < 0]
            for i in shiftRequired.index:
                neededShift = -1 * shiftRequired['Slack'].loc[i]
                temp = processedSample[processedSample['Slack'] > 0]
                if len(temp) == 0:
                    raise ValueError("More Samples were Requested than Sampling Units")
                newProposal = rd.choices(temp['Unit'].tolist(),k = neededShift)


                processedSample.loc[processedSample['Unit'] == shiftRequired['Unit'].loc[i],'Samples'] -= neededShift 
                processedSample.loc[processedSample['Unit'] == shiftRequired['Unit'].loc[i],'Slack'] = 0
                for element in newProposal:
                    processedSample.loc[processedSample['Unit'] == element,'Samples'] += 1 
                    processedSample.loc[processedSample['Unit'] == element,'Slack'] -= 1 

        counter = 0
        for i in processedSample.index:
            tempFrame = floorFrame.loc[floorFrame['samplingUnit'] == processedSample["Unit"].loc[i],:].sample(n = processedSample["Samples"].loc[i],random_state = seed)
            seed += 1 
            sampleFrame = sampleFrame.append(tempFrame,ignore_index = True)
            
    return sampleFrame


def validateProposedSample(proposal,originalFrame,target):
    allUnits = np.array(list(set(proposal)))

    #Create Info Frame
    processedData = pd.DataFrame({"Unit": allUnits,
                                  "Samples": [0] * len(allUnits),
                                  "Slack": [-1] * len(allUnits)})

    for unit in allUnits:
        processedData.loc[processedData["Unit"] == unit,"Samples"] = sum(proposal == unit)
        processedData.loc[processedData["Unit"] == unit,"Slack"] = sum(originalFrame[target] == unit) - processedData.loc[processedData["Unit"] == unit,"Samples"]

    return processedData


def getDormStructure(processedDormTable):
    #Create a pandas table to store the data
    dormStructure = pd.DataFrame({'dorm':np.array([],dtype = 'U'),
                                  'floor':np.array([],dtype = int),
                                  'bathroomType':np.array([],dtype = int),
                                  'units':np.array([],dtype = int)})

    for dorm in set(processedDormTable['Building']):
        for floor in set(processedDormTable.loc[processedDormTable['Building'] == dorm,'Floor']):
            currentFloorData = processedDormTable.loc[(processedDormTable['Building'] == dorm) & (processedDormTable['Floor'] == floor)]
            if (currentFloorData['bathroom'].array == None).all():
                for room in set(currentFloorData['room']):
                    configuration = str(list(currentFloorData.loc[currentFloorData['room'] == room,'occupancy']))
                    if configuration in dormStructure.loc[(dormStructure['dorm'] == dorm) & (dormStructure['floor'] == floor),'bathroomType'].array:
                        dormStructure.loc[(dormStructure['dorm'] == dorm) & (dormStructure['floor'] == floor) & (dormStructure['bathroomType'] == configuration),'units'] += 1
                    else:
                        dormStructure = dormStructure.append(pd.DataFrame({'dorm':np.array([dorm]),
                                                                           'floor':np.array([floor]),
                                                                           'bathroomType':np.array([configuration]),
                                                                           'units':np.array([1])}),ignore_index = True)
            elif (currentFloorData['bathroom'].array == None).any():
                for bathroom in set(currentFloorData['bathroom']):
                    if bathroom:
                        bathroomConfig = currentFloorData.loc[currentFloorData['bathroom'] == bathroom,'occupancy']
                        configuration = list(bathroomConfig)
                        configuration.sort()
                        configuration = str(configuration)
                        if configuration in dormStructure.loc[(dormStructure['dorm'] == dorm) & (dormStructure['floor'] == floor),'bathroomType'].array:
                            dormStructure.loc[(dormStructure['dorm'] == dorm) & (dormStructure['floor'] == floor) & (dormStructure['bathroomType'] == configuration),'units'] += 1
                        else:
                            dormStructure = dormStructure.append(pd.DataFrame({'dorm':np.array([dorm]),
                                                                               'floor':np.array([floor]),
                                                                               'bathroomType':np.array([configuration]),
                                                                               'units':np.array([1])}),ignore_index = True)
                    else:
                        for room in set(currentFloorData.loc[currentFloorData['bathroom'].array == None,'room']):
                            configuration = str(list(currentFloorData.loc[currentFloorData['room'] == room,'occupancy']))
                            if configuration in dormStructure.loc[(dormStructure['dorm'] == dorm) & (dormStructure['floor'] == floor),'bathroomType'].array:
                               dormStructure.loc[(dormStructure['dorm'] == dorm) & (dormStructure['floor'] == floor) & (dormStructure['bathroomType'] == configuration),'units'] += 1
                            else:
                               dormStructure = dormStructure.append(pd.DataFrame({'dorm':np.array([dorm]),
                                                                                  'floor':np.array([floor]),
                                                                                  'bathroomType':np.array([configuration]),
                                                                                  'units':np.array([1])}),ignore_index = True)
            else:
                for bathroom in set(currentFloorData['bathroom']):
                    bathroomConfig = currentFloorData.loc[currentFloorData['bathroom'] == bathroom,'occupancy']
                    configuration = list(bathroomConfig)
                    configuration.sort()
                    configuration = str(configuration)
                    if configuration in dormStructure.loc[(dormStructure['dorm'] == dorm) & (dormStructure['floor'] == floor),'bathroomType'].array:
                        dormStructure.loc[(dormStructure['dorm'] == dorm) & (dormStructure['floor'] == floor) & (dormStructure['bathroomType'] == configuration),'units'] += 1
                    else:
                        dormStructure = dormStructure.append(pd.DataFrame({'dorm':np.array([dorm]),
                                                                           'floor':np.array([floor]),
                                                                           'bathroomType':np.array([configuration]),
                                                                           'units':np.array([1])}),ignore_index = True)
    return dormStructure
```
