## Supplementary material for "Monitoring and responding to emerging infectious diseases in a university setting: A case study using COVID-19": S2 Supporting Information: README.rst

This directory contains three Python scripts and one R script that replicate the analyses in Soda, Chen, Feinn, and Hill. You must have an installation 
of base Covasim and our extensions to the model installed on your computer before most analyses can be run. You may download the extended version of the 
model from GitHub (https://github.com/kjamessoda/covasim.git). The model version used in the paper is contained in the F20ModelAndTesting branch.

################################
QUCovasimAnalyses_PLOSReady.py
################################
This script can implement any of the 28 scenarios/sub-scenarios discussed in the paper. To run a scenario, specify the scenario's name (see below) as
the value of the variable scenario. In addition to the standard Covasim model outputs, the script will also save the simulated time series of cumulative
infections, cumulative diagnosed cases, and cumulative tests used to a CSV file. You may specify these files' paths (including their names) using the
variables seriesPath (for the cumulative infections), diagnosedFilePath (for the cumulative diagnosed cases), and testsFilePath (for the cumulative
tests). You may also elect to suppress the creation of these files by changing writeInfectionToFile to False.

The possible values of scenario are listed below. Scenarios are grouped based on which component of the paper it relates to.

---------------
Main Scenarios
---------------
*"R0_1.5_Contacts_8"
*"R0_1.5_Contacts_10"
*"R0_2.5_Contacts_8"
*"R0_2.5_Contacts_10"

In the values above, the number after R0 refers to the basic reproductive number for the corresponding scenario, and the number after Contacts refers to
the average number of contacts per day each agent has in the corresponding scenario.

----------------------------------------------------------
Sensitivity Analysis for the Rate of Imported Infections
----------------------------------------------------------
*"R0_1.5_Contacts_8_HalfImport"
*"R0_1.5_Contacts_8_DoubleImport"
*"R0_1.5_Contacts_10_HalfImport"
*"R0_1.5_Contacts_10_DoubleImport"
*"R0_2.5_Contacts_8_HalfImport"
*"R0_2.5_Contacts_8_DoubleImport"
*"R0_2.5_Contacts_10_HalfImport"
*"R0_2.5_Contacts_10_DoubleImport"

See the description in Main Scenarios for how to interpret the numbers after R0 and Contacts. Values that end with HalfImport correspond to scenarios
that have half the rate of imported infections relative to its corresponding main scenario, and values that end with DoubleImport correspond to scenarios
that have twice the rate of imported infections relative to its corresponding main scenario.

-----------------------------------
Comparison of Sampling Strategies
-----------------------------------
*"R0_1.5_Contacts_8_SRS"
*"R0_1.5_Contacts_8_100percent"
*"R0_1.5_Contacts_10_SRS"
*"R0_1.5_Contacts_10_100percent"
*"R0_2.5_Contacts_8_SRS"
*"R0_2.5_Contacts_8_100percent"
*"R0_2.5_Contacts_10_SRS"
*"R0_2.5_Contacts_10_100percent"

See the description in Main Scenarios for how to interpret the numbers after R0 and Contacts. Values that end with SRS correspond to scenarios
that use simple random sampling to implement surveillance testing, and values that end with 100percent correspond to scenarios that sample every agent to
implement surveillance testing.

-------------------
Holdout Scenarios
-------------------
*"Holdout_AllInterventions_Targeted"
*"Holdout_AllInterventions_SRS"
*"Holdout_DelayedSymptomTesting_Targeted
*"Holdout_DelayedSymptomTesting_SRS"
*"Holdout_noSymptomTesting_Targeted"
*"Holdout_noSymptomTesting_SRS"
*"Holdout_noContactedTracing_Targeted
*"Holdout_noContactedTracing_SRS"

Values that contain AllInterventions implement all four interventions discussed in the paper, but unlike the main scenarios, do not include any 
non-residential students. Values that contain DelayedSymptomTesting correspond to the delayed-symptomatic-testing scenario. Values that contain 
noSymptomTesting correspond to the no-symptomatic-testing scenario. Values that contain noContactTracing correspond to the no-contact-tracing scenario.
Values that end in Targeted use a targeted sampling design to implement surveillance testing, whereas values that end in SRS correspond to scenarios
that use simple random sampling to implement surveillance testing.

################################
QUDormsLegacy_20200904_208.py
################################
This Python script contains a helper function that constructs the Dorm objects used in QUCovasimAnalyses_PLOSReady.py.

################################
estimateScalingRelationships.py
################################
This Python script estimates the proportional scaling relationship between the final cumulative infections and the rate of imported infections under each
of the main scenarios. The script relies on the files provided in the sensitivityFiles subdirectory. Each file in this subdirectory contains a simulated
time series of cumulative infections for one of the main scenarios or sensitivity analysis scenarios. These same files can also be created using 
QUCovasimAnalyses_PLOSReady.py but are provided in the sensitivityFiles subdirectory for convenience.

#######################################
betaAndRateOfImportationCalculations.R
#######################################
This R script contains functions and code to estimate the base probability of transmission when an infectious individual contacts a susceptible 
individual and the rate of imported infections under the four main scenarios.
